# Social and structural determinants of injection drug use-associated bacterial and fungal infections: a qualitative systematic review and thematic synthesis

**DOI:** 10.1101/2022.10.02.22280620

**Authors:** Thomas D. Brothers, Matthew Bonn, Dan Lewer, Emilie Comeau, Inhwa Kim, Duncan Webster, Andrew Hayward, Magdalena Harris

**Author notes:** Address correspondence to; (TDB).

## Abstract

**Background:** Injection drug use-associated bacterial and fungal infections are increasingly common, and social contexts shape individuals’ injecting practices and treatment experiences. We sought to synthesize qualitative studies of social-structural factors influencing incidence and treatment of injecting-related infections.

**Methods:** We searched PubMed, EMBASE, Scopus, CINAHL, and PsycINFO from January 1, 2000, to February 18, 2021. Informed by Rhodes’ “risk environment” framework, we performed thematic synthesis in three stages: (1) line-by-line coding; (2) organizing codes into descriptive themes, reflecting interpretations of study authors; (3) consolidating descriptive themes into conceptual categories to identify higher-order analytic themes.

**Results:** We screened 4,841 abstracts and included 26 qualitative studies on experiences of injecting-related bacterial and fungal infections. We identified six descriptive themes organized into two analytic themes. The first analytic theme, *social production of risk*, considered macro-environmental influences. Four descriptive themes highlighted pathways through which this occurs: (1) *unregulated drug supply*, leading to poor drug quality and solubility; (2) *unsafe spaces*, influenced by policing practices and insecure housing; (3) *health care policies and practices*, leading to negative experiences that discourage access to care; and (4) *harm reduction programs*, including structural barriers to effective service provision. The second analytic theme, *practices of care among people who inject drugs*, addresses protective strategies that people who inject drugs employ within infection risk environments. Associated descriptive themes were: (5) *mutual care*, including assisted-injecting and sharing sterile equipment; and (6) *self-care*, including vein health and self-treatment. Within constraining risk environments, some protective strategies for bacterial infections precipitated other health risks (e.g., HIV transmission).

**Conclusions:** Injecting-related bacterial and fungal infections are shaped by modifiable social-structural factors, including unregulated drug quality, criminalization, insufficient housing, limited harm reduction services, and harmful health care practices. Enabling environment interventions that address these factors could further empower people who inject drugs to protect themselves and their community.

## INTRODUCTION

Injection drug use-associated bacterial and fungal infections (e.g. skin and soft-tissue infections, endocarditis, epidural abscess) cause significant morbidity and mortality among people who inject drugs (PWID).^1–6^ The incidence of hospitalizations for severe injecting-related infections is increasing in Australia,^7^ Canada,^2,8^ the United Kingdom (UK),^9^ and the United States of America (USA).^10–14^

Efforts to prevent injecting-related bacterial and fungal infections have focused on individual-level behavioural interventions,^15,16^ including education on hand-washing before drug preparation,^17^ skin-cleaning before injecting,^18^ and avoiding subcutaneous/intramuscular injecting.^19^ While individual-level interventions may be helpful for PWID who can adopt these practices, evaluations of these interventions have shown mixed results^20–22^ and the incidence of injecting-related infections continues to rise.

Risk for injecting-related bacterial and fungal infections likely reflects contributions of multiple factors external to PWID that enable and/or constrain their behaviour and influence health outcomes.^23–26^ Identifying, measuring, and ameliorating such social-structural factors has informed clinical and public health responses to other drug-related harms, including HIV,^27–29^ hepatitis C virus (HCV),^30^ and overdose.^31,32^ Understanding the influence of social context on health can broaden awareness of the causes of illness^33^ and inform more appropriate prevention and treatment interventions.^29,34,35^

### Objectives

To understand social-structural determinants of injecting-related bacterial and fungal infections and to identify opportunities for potential intervention, we aimed to: (1) systematically review qualitative studies on experiences of injecting-related infections, and (2) synthesize analyses of social-structural factors influencing risk for injecting-related infections, their treatment, and subsequent health outcomes.

## METHODS

Before conducting the search, we published our protocol^1^ and registered with PROSPERO (CRD42021231411). We modified our original protocol after our search and full-text review. Our protocol specified a “mixed studies review” of quantitative, qualitative, and mixed-methods studies.^38,39^ As we identified more and richer qualitative sources than anticipated, we decided to consider qualitative and quantitative data separately. Here, we report the qualitative systematic review and thematic synthesis. Quantitative results will be reported separately. This manuscript follows PRISMA guidelines^36^ and the ENTREQ statement^37^ on qualitative systematic reviews.

### Conceptual model and framework

The “risk environment”, as developed by Rhodes and others,^27,29,40,41^ is a socio-ecological model describing how macro-environmental (e.g., criminalization; racism) and micro-environmental (e.g., local availability of needle and syringe programs) factors interplay to influence health practices and outcomes.^42^ The risk environment model encourages thinking about how people interact with and modify constraining environments (e.g., drug users’ unions organizing to repeal laws banning supervised consumption sites).^43^ Collins and colleagues recently extended the risk environment to incorporate intersectionality, considering how social-structural factors affect PWID differently depending on other social identities and their locations within power hierarchies, including race and gender.^42^

Injecting-related bacterial and fungal infections occur through introducing bacteria or fungi into sterile tissues (often from commensal organisms living on the skin) and are precipitated by particulate matter that damages blood vessels, lymphatics, and heart valves.^35,44^ To conceptualise how the risk environment affects injecting-related infections at different times, we developed a framework (see Figure 1) illustrating a pathway from (a) drug acquisition (e.g. solubility of drugs); (b) drug preparation (e.g. using sterile water to dissolve drugs); (c) drug injection (e.g. accessing veins to avoid intramuscular injecting); (d) development of and care for superficial infections (e.g. self-treatment; primary care); (e) development of and care for severe infections (e.g. hospitalization); and (f) outcomes after infections (e.g. access to follow-up care).^1^ Not all PWID would progress through all stages; some PWID do not develop infections and many who develop infections never access treatment.

**Figure 1.**
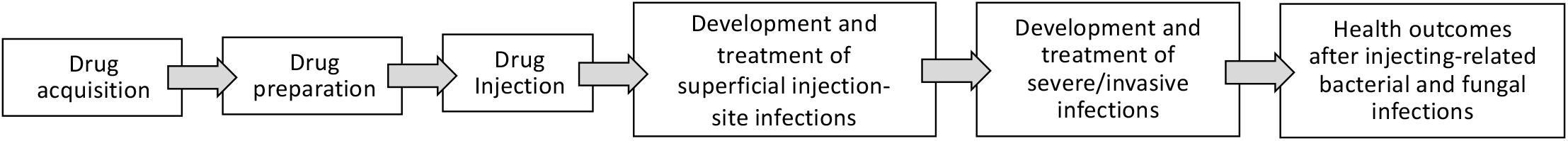
Illustrative schematic of pathway model to conceptualize how the risk environment shapes risk for injecting-related bacterial and fungal infections at different moments. Macro-environmental, micro-environmental, and individual-level factors interplay to influence risk at each moment.

### Eligibility criteria

A full description is in our published protocol.^1^ Briefly, we included articles in peer-reviewed journals reporting qualitative studies. We followed the Population, Exposures, Outcomes approach to eligibility criteria.^45^ The population was PWID (i.e., people injecting any psychoactive substance; excluding people only injecting performance-enhancing or gender-affirming hormones). Exposures were any social or environmental factors that may affect risk of infections, such as housing, service availability, or policing practices. Outcomes included incidence, treatment, or outcomes of injecting-related bacterial and fungal infections. Eligible studies were published in English or French between January 1, 2000, and February 18, 2021 (to capture contemporary research more likely to inform policy and clinical practice).

### Information sources and search strategy

We searched PubMed, EMBASE, Scopus, CINAHL, and PsycINFO databases. We developed the search strategy in consultation with a librarian (see Supplementary Table S1 for full search strategy). We supplemented searches with backward and forward citation chaining and with other studies known to the review team (which includes people with lived/past and living/current experience of injection drug use, researchers, and clinicians caring for PWID).

### Data management and reference selection

We uploaded titles/abstracts into Covidence software, where they were automatically de-duplicated. Two reviewers (TDB and either MB, DL, EC, or IK) independently screened titles/abstracts, resolving discrepancies through consensus. We obtained full-text reports for sources that passed screening, and one reviewer (TDB) assessed full-text reports.

### Quality assessment

We used the Mixed Methods Appraisal Tool (MMAT), which is a validated and commonly-used appraisal tool for mixed studies reviews.^46,46,47^ TDB and EC independently appraised each study, resolving discrepancies through discussion. For data synthesis, we included studies meeting both MMAT screening questions: “Are there clear research questions?” and “Do the collected data allow to address the research questions?”

### Data synthesis

Following Thomas and Harden,^48–51^ thematic synthesis comprises three stages: (1) line-by-line open coding; (2) organizing codes into descriptive themes reflecting content of studies and study authors’ interpretations; (3) translating descriptive themes and associated codes across studies to generate analytic themes. Coding and generation of descriptive themes focuses on study authors’ analysis and interpretation because reviewers do not have full knowledge of the original study data.^48,49^

First, TDB (physician and PhD student with qualitative methods training) familiarized himself with the included studies. Next, MB (researcher with lived/living experience of injecting-related infections and a drug policy activist) and TDB independently performed line-by-line coding on the same three purposefully selected, data rich sources.^52–54^ They compared and contrasted codes and revised them in an iterative, deductive-inductive process, informed by the risk environment model.

The whole review team met to provide feedback on these candidate codes: DL (public health specialist), EC and IK (medical students), DW (infectious diseases and addiction medicine physician), AK (infectious disease epidemiologist), and MH (health sociologist with lived experience of injection drug use). TDB coded the remaining papers over several rounds, including adding and revising new candidate codes after discussing with the team at meetings and through collaborative online writing.

TDB developed descriptive themes by comparing and contrasting codes across studies, seeking to organize codes into related social-structural categories and proposed them to the team for feedback. TDB then consolidated descriptive themes into conceptual categories to generate analytic themes that were finalized over several iterations and team meetings.

## RESULTS

Following de-duplication, we screened 4,841 titles/abstracts and evaluated 631 full-text reports. After considering 16 additional reports identified outside the search, we identified 151 eligible studies (quantitative, qualitative, and mixed-methods) for our “mixed studies” review. Here, we report on the 26 studies with qualitative data and analysis (19 qualitative-only and seven mixed-methods). See Figure 2 for PRISMA diagram. All 26 qualitative studies met our quality criteria for inclusion (see Supplementary Table S2 for full MMAT results).

**Figure 2.**
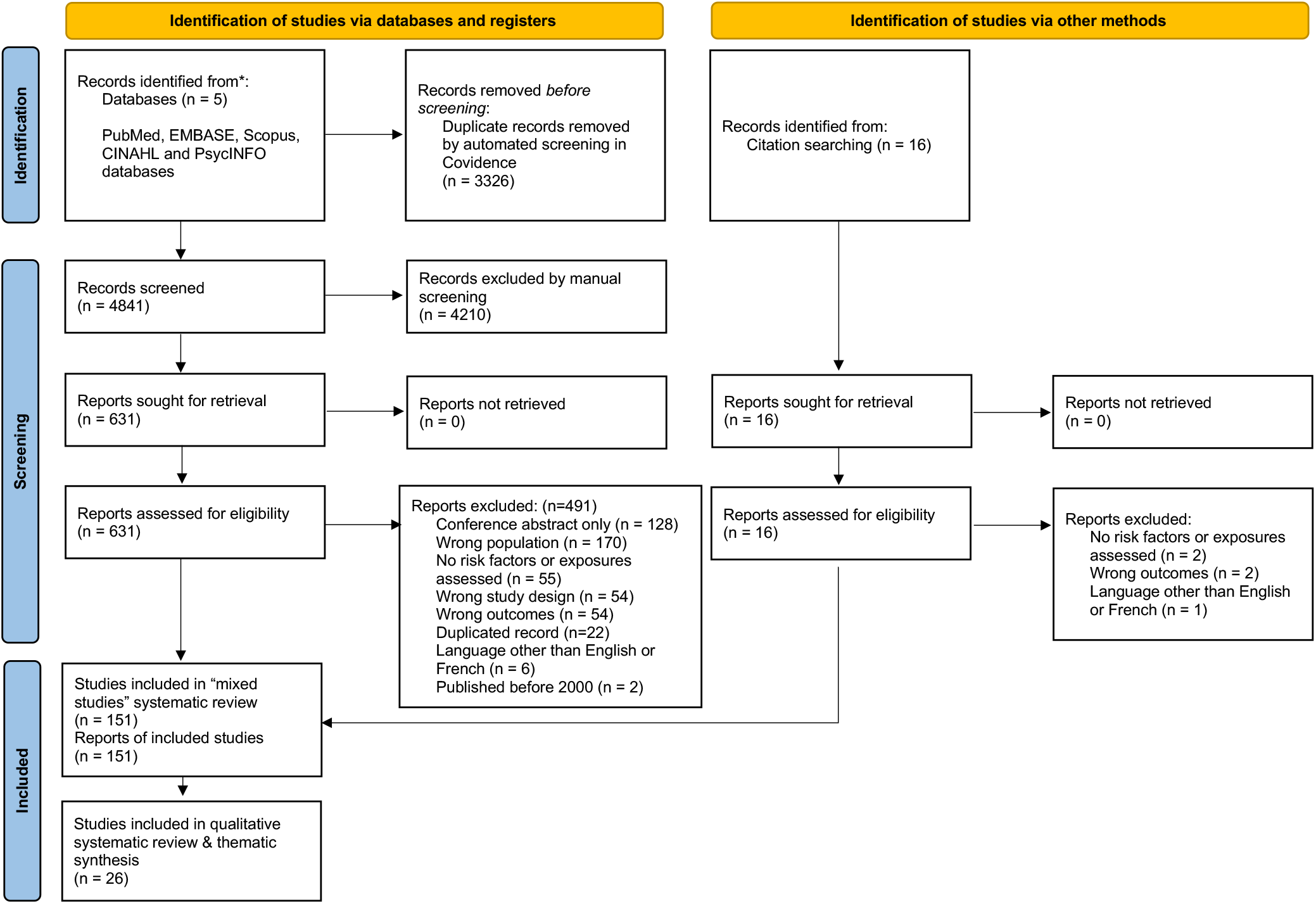
PRISMA flow diagram of included studies in systematic review and thematic synthesis of social and structural determinants of injection drug use-associated bacterial and fungal infections.

### Study characteristics

See Table 1 for summaries of individual studies. The majority (n=20 studies) were conducted in North America. Qualitative data came from individual interviews (n=23), observation/ethnography (n=4), and focus groups (n=2). Studies included experiences of injecting-related skin and soft-tissue infections (n=22), endocarditis (n=7), bacteremia (n=3), and osteomyelitis (n=2). All 26 studies included bacterial infections, and only one study^55^ included fungal infections (candidemia and fungal ophthalmitis).

**Table 1.** Summary of included studies.

| Study | Country | Qualitative study design | Sample | Focus of interviews/analysis | Conceptual or explanatory model(s) | Summary of findings |
| --- | --- | --- | --- | --- | --- | --- |
| Bearnot 2019<sup>64</sup> | USA | Individual interviews<br>Grounded theory | 11 PWID with opioid injection-associated endocarditis<br>55% women; median age 38 years; 55% with “unstable housing” | Experiences of endocarditis care | None specified | Poor health outcomes among PWID with opioid injection-associated endocarditis are caused by stigma, delays or discontinuity of care, social and medical comorbidities, perceptions of addiction as a chronic and relapsing disease, and prolonged hospitalizations.. |
| Bearnot 2020<sup>63</sup> | USA | Secondary analysis of interview data from Bearnot 2019<sup>64</sup><br>Journey mapping analysis<br>Grounded theory | Same as Bearnot 2019 | Patterns of care for endocarditis | None specified | People with opioid injection-associated endocarditis left care before medically advised because of poor care experiences, including undertreatment of withdrawal and pain and discrimination from clinicians. Following hospitalization, participants commonly engaged in outpatient addiction treatment and follow-up endocarditis care. Leaving outpatient addiction treatment often preceded rehospitalizations with recurrent infections. |
| Bodkin 2015<sup>70</sup> | Canada | Individual interviews<br>Qualitative descriptive analysis | 14 PWID recruited from an outreach program for sex workers in London, Ontario | Access to health care among PWID who do sex work | None specified | Sex workers who inject drugs avoided primary care and emergency department treatment of injecting-related infections because of experiences of stigmatization and |
|  |  |  | 100% women; age range 23-49 years; housing status not reported |  |  | criminalization. Participants experienced involuntary discharge from hospital and received suboptimal oral antibiotics because of abstinence-requiring policies in hospital. |
| Bourgois 2006<sup>59</sup> | USA | Mixed methods<br>Participant-observation ethnography (field notes, interviews, photographs) | Sample not specifically described, but includes African American and white men who inject heroin in San Francisco, California | How social-structural determinants interface with drug consumption practices and survival strategies among African American and white PWID | "...a social science theoretical understanding of the link between large-scale power relations and individual risky practices that shape the spread of blood-borne disease among injectors." | Higher rates of abscesses among white PWID compared to African American PWID reflect socially produced differences in norms between racial/ethnic groups, including social acceptability of subcutaneous injecting among white PWID. Conversely, police are more likely to repeatedly search and confiscate sterile injecting equipment from African American PWID. |
| Case 2008<sup>75</sup> | Mexico | Individual interviews<br>Thematic content analysis | 43 PWID recruited through street-based outreach, shooting galleries, and drug treatment programs in Tijuana (n=20) and Ciudad Juárez (n=23)<br>42% women; median age 30; 30% lived or slept | Injection methamphetamine use in two Mexican border cities | Structural vulnerability | Greater availability of methamphetamine in Tijuana is associated with widespread use, compared to Ciudad Juárez. Injecting methamphetamine is perceived to be associated with increased risk of injecting-site abscesses, described more commonly in Tijuana. |
|  |  |  | "on streets" in past six months |  |  |  |
| Colwill 2021<sup>69</sup> | USA | Individual interviews<br>Grounded theory | 11 PWID undergoing surgical evaluation for injecting-related endocarditis<br>45% women; mean age 31; 9% "Homeless" | Experiences of endocarditis | "PWID with Endocarditis Cyclical Experiences (PEaCE) model" | PWID with endocarditis avoid health care because of stigmatizing and discriminatory experiences. The experience of endocarditis motivated some participants to enter addiction treatment and pursue abstinence. |
| Dunleavy 2019<sup>52</sup> | Scotland | Individual interviews<br>Framework analysis | 22 PWID who had experienced a skin and soft-tissue infection within past year, recruited from needle and syringe program (Glasgow; n=14) or drug treatment service (Edinburgh; n=8)<br>32% women; median 36 years; 59% experienced homelessness during the past six months | Experiences of skin and soft-tissue infections | Rhodes' risk environment | Stigma associated with skin and soft-tissue infections motivated some participants to try to reduce risks, including use of sterile equipment and safer injecting techniques. No participants had learned about infections and associated risks from harm reduction programs. Social and environmental factors contributing to infection risk included insufficient access to sterile injecting equipment, caustic adulterants in the local drug supply, and a lack of hygienic spaces to prepare and consume drugs. |
| Epele 2002<sup>60</sup> | USA | Individual interviews, observations, and | 35 PWID recruited through syringe service | Risk conditions and care practices related to HIV | Political economy of health | Some women who inject drugs rely on injecting assistance from others to avoid intramuscular injection and |
|  |  | participation in everyday life settings<br>Analysis approach not specified | program and snowball sampling<br>71% women; mean age of women was 34; all “do not consider themselves as homeless” | among Latino women | Biopower | associated abscesses and scars, because these lead to loss of social status and negatively affect relationships and potential income generation through sex work. Participants recognized that assisted-injecting increases risks for HIV infection. Injection assistance is provided by friends, romantic or sexual partners, and paid “hit doctors”. |
| Gilbert 2019<sup>71</sup> | USA | Secondary analysis of interview data from Summers 2018<sup>72</sup><br>Thematic analysis | Same sample as qualitative sample in Summers 2018<sup>72</sup><br>12 clients of a syringe services program in Boston (n=6) and Sacramento (n=6)<br>25% women; median 46 years; housing status not reported | Experience of skin and soft-tissue infections | Health belief model (HBM) of health-seeking behaviors<br>Conceptual Model of Medical Care Avoidance | PWID had good knowledge about skin infections and avoided formal health care due to traumatic experiences, discrimination, and unnecessarily painful procedures. Participants described multiple strategies for prevention and treatment of injecting-related infections including hydration, topical applications, non-prescribed antibiotics, and incision and drainage by non-medical providers. |
| Harris RE 2018a<sup>53</sup> | USA | Individual interviews<br>Analysis approach not specified | 19 clients of a syringe services program in Philadelphia<br>53% women; median age 39 [27-59 years]; | Experiences of skin and soft-tissue infections | None specified | PWID described good knowledge about risks of injecting-related skin infections, but were prevented from using hygienic techniques as they lacked of safe places to use drugs. Participants therefore injected abandoned buildings or outdoors, with inadequate lighting, or fear of assault |
|  |  |  | housing status not reported |  |  | or arrest, leading to drug contamination and intramuscular injection. Participants tended to avoid medical care for injecting-related infections due to prior negative healthcare experiences, including stigma and inadequate treatment of withdrawal and pain. Some participants described self-treatment of infections, including increased drug use for pain control and performing incision and drainage on themselves. |
| Harris RE 2018b<sup>58</sup> | USA | Individual interviews<br>Thematic analysis | Same sample as Harris RE 2018a<sup>53</sup>.<br>19 clients of a syringe services program in Philadelphia.<br>53% women; median 39 years; housing status not reported | Perceptions of safe injecting facilities | Rhodes' risk environment | PWID described commonly being forced to inject in public spaces, which led them to rush and inject intramuscularly or subcutaneously for fear of assault or arrest. Participants supported the idea of a supervised injection site to reduce these risks and inject more safely. Participants with stable housing preferred to inject at home and described that this reduced risks of injecting-related infections due to less fear of assault or arrest, adequate light and heat, running water, and space to store sterile injecting equipment. |
| Harris M 2019<sup>55</sup> | England | Mixed methods<br>Individual interviews<br>Constructivist grounded theory | 31 PWID recruited through drug treatment services, homeless hostels, and day centres | Use of acidifiers | Rhodes' risk environment | Excessive acidifier use in drug preparation for injection is common and contributes to venous damage and risk for bacterial infections. Some participants determined the amount of acidifier to use through expert practice (e.g., visual cue of solution |
|  |  |  | across London, UK<br>29% women; mean age 43 years; housing status not reported |  |  | clarity) and others through external factors (e.g., using one whole packet of acid, even if that is excessive and causes pain and injury). Some participants decreased acidifier use over time, in response to new information or pain/injury. The authors infer a need to revisit design and distribution of acidifiers within harm reduction programs. |
| Harris M 2020a<sup>61</sup> | England | Individual interviews<br>Constructivist grounded theory | 36 PWID, recruited through specialist drug services, homeless hostels, and day centres across London, UK.<br>12% women; mean 46 years; 64% unstably housed in past 12 months. | Experiences of injecting-related injuries and infections | Everyday violence<br>Structural violence<br>Cultural safety | Engagement with the medical system (including for injecting-related infections) is a “last resort”; often participants delayed as long as possible to the point that they were critically ill. Participants avoided or delayed accessing medical care for their own protection, including because of experiences of discrimination and undertreated withdrawal and pain; one participant specifically worried of stigma against mothers who use drugs and associated risks of child apprehension. Participants described leaving hospital prematurely and self-treating wounds instead. |
| Harris M 2020b<sup>56</sup> | England | Mixed methods<br>Individual interviews<br>Constructivist grounded theory | 32 PWID, recruited through specialist drug services, homeless hostels, and day centres | Water for preparing injecting solutions | Rhodes’ risk environment | Environmental constraints to sourcing sterile water for injection preparation (and staying hydrated to promote vein health) include lack of housing, public washrooms, or sterile water from harm reduction programs. When injecting in public places, fear of arrest |
|  |  |  | across London, UK.<br>31% women; mean age 43 years; 94% had experienced homelessness |  |  | would lead people to rush their preparation and inject as fast as possible. As a result, participants described using more readily available but unsafe alternative water sources including puddle water, toilet cistern water, whisky, cola soda, and saliva to prepare injections, which were associated with bacterial infections. Participants described several strategies to promote health and safety despite these environmental constraints, including filtering water through alcohol swabs or asking passers-by for bottled water. |
| Jafari 2015<sup>74</sup> | Canada | Mixed methods<br>Individual interviews; direct observation and field notes<br>Narrative analysis | 8 PWID who were clients at a harm reduction-oriented medical respite program<br>Gender, age, and current housing status not reported | Experiences of care for injecting-related infections | None specified | Participants described past experiences of leaving hospital before completion of their medical treatment because of judgmental and stigmatizing care. Clients with severe injecting-related infections who were being cared for at a harm reduction-oriented medical respite describe receiving less judgmental and stigmatizing care compared to their experience in acute care hospitals. |
| Krüsi 2009<sup>65</sup> | Canada | Individual interviews<br>Thematic analysis | 22 PWID, recruited as clients at an HIV-focused residential and outpatient care facility in Vancouver | Use of a supervised injection site integrated within a community-based HIV care facility | Rhodes' risk environment | Participants accessing the supervised injection site found it a uniquely valuable setting to receive education on (and to implement) safer drug preparation and injecting techniques, which they attributed to reduced frequency of abscesses. When they did not have access to the supervised |
|  |  |  | 32% women;<br>mean 44 years;<br>housing status<br>not reported |  |  | injecting facility, participants described rushing their drug preparation and injection out of fear, including not using water to dissolve their heroin sufficiently. |
| Mars<br>2016<sup>57</sup> | USA | Ethnography and<br>individual interviews<br>Grounded theory | 41 PWID<br>recruited during<br>ethnographic<br>insertion in drug<br>using community<br>and with snowball<br>sampling in<br>Philadelphia<br>(n=22) and San<br>Francisco (n=19)<br>49% women; age<br>unknown;<br>homelessness<br>“common”. | Comparing<br>perspectives of<br>PWID in two<br>different heroin<br>markets. | Rhodes’ risk<br>environment | In San Francisco, where heroin was mostly in “tar” form, participants attributed abscesses to the characteristics of tar heroin including poor solubility. In Philadelphia, where more-soluble power heroin as well as cocaine was widely available, participants attributed abscesses to missing veins (i.e., injecting subcutaneously or intramuscularly) and when injecting cocaine. The authors attribute regional differences in abscess risk to geopolitical forces that have segmented the U.S. heroin market. |
| McNeil<br>2014<sup>67</sup> | Canada | Individual interviews<br>Thematic analysis | 30 PWID who had<br>experienced<br>hospital discharge<br>against medical<br>advice within the<br>prior two years,<br>recruited from<br>within a<br>prospective<br>cohort study in<br>Vancouver<br>43% women;<br>mean 45 years; | Hospital care<br>experiences | Rhodes’ risk<br>environment<br>Social violence | Participants left hospital prematurely (before the completion of their recommended treatment) because of inadequate pain and withdrawal management, and because of discriminatory, stigmatizing, and racist care experiences. These were influenced by hospital policies, written and unwritten. |
|  |  |  | 27% staying in emergency shelters or unhoused |  |  |  |
| Meyer 2020<sup>73</sup> | Kyrgyzstan | Individual interviews<br>Content analysis | 11 PWID who were incarcerated and injected diphenhydramine<br>10% women; average age not reported; all currently incarcerated | Diphenhydramine injecting in Kyrgyz prisons | Rhodes' risk environment | Participants attributed severe skin infections to injecting diphenhydramine while incarcerated, particularly in comparison to injecting heroin. Infectious risks associated with diphenhydramine were influenced by the denial of access to the prison's needle and syringe program to people taking methadone (which was common among people injecting diphenhydramine) and stigmatization and punishment of diphenhydramine users in the prison (which led people to delay seeking care for skin infections). |
| Paquette 2018<sup>68</sup> | USA | Individual interviews<br>"Mixed inductive and deductive approach" | 46 PWID who attended syringe service programs or health services in Fresno, California (n=22) or community services agencies or street-based recruitment in Kern, California (n=24)<br>37% women; mean 39 years; | Stigmatizing health care experiences | Rhodes' risk environment | Participants described delaying or avoiding medical care ("until it was absolutely necessary") for injecting-related infections because of previous stigmatizing experiences. Instead, some people treat their own infections. Participants in rural areas also described feeling as if they could not attend their local harm reduction program for sterile injecting equipment as this would "out" them as a drug user to the small community. |
|  |  |  | housing status not reported |  |  |  |
| Phillips 2013<sup>54</sup> | USA | Mixed methods<br>Focus group interviews<br>Qualitative analysis approach not specified | 32 PWID recruited through street outreach in Denver, Colorado<br>50% women; mean age 50 years; housing status not reported | Perspectives on injecting-related bacterial infections, to inform development of a behaviour change intervention | Information-Motivation-Behavioral Skills model | Most participants had experienced injecting-related bacterial infections. PWID attributed increased risk of infections to poor quality (or adulterated or contaminated) unregulated drugs, including tar heroin (compared to powder heroin or pharmaceutical opioid tablets); to injecting intramuscularly or subcutaneously; to reusing needles; and to not cleaning skin. Barriers to practicing safer drug preparation and injecting included lack of access to sterile equipment (influenced by a “paraphernalia law” that prohibited carrying a hypodermic needle without “proof of medical need”).<br>Participants described delaying or avoiding medical care for infections due to negative health care experiences. |
| Pollini 2010<sup>76</sup> | Mexico | Mixed methods<br>Focus groups<br>Grounded theory | 47 PWID invited from among participants in a cohort study that used respondent-driven sampling<br>14% women; age not reported; | Barriers to sterile syringe access, including purchase from pharmacies | Rhodes’ risk environment | Participants described many challenges in accessing sterile needles and syringes via purchasing at local pharmacies, including discrimination from pharmacists and pharmacists disclosing fear of “trouble with police” (despite syringe sales being legal). This led to syringe reuse being common practice. Participants did not spontaneously attribute risks for |
|  |  |  | housing status not reported |  |  | abscesses to needle and syringe reuse, until asked by a focus group facilitator. |
| Pollini 2021<sup>62</sup> | USA | Individual interviews<br>Thematic analysis | 20 PWID recruited through provider referral, street-based recruitment and snowball sampling<br>45% women; median age 26; housing status not reported | Scarcity of sterile needles and syringes in a rural environment | Rhodes' risk environment | Scarcity of sterile needle and syringes led PWID to share and re-use syringes. Factors limiting sterile syringe access included pharmacies refusing to sell them or requiring an ID, and a state "drug paraphernalia" law that criminalizes possession of syringes. Participants would travel out-of-state to pharmacies that would sell syringes, but police were aware of this and would stop and search cars with out-of-state license plates after visiting a pharmacy. One participant obtained sterile syringes from a family member with diabetes and distributed them to people in her community who inject drugs. |
| Sheard 2008<sup>77</sup> | England | Individual interviews<br>Grounded theory | 45 women who inject drugs recruited among clients of needle and syringe programs and addiction treatment programs in semirural North Nottinghamshire and urban Leeds, and through snowball sampling of | Assistance with injecting among women. | None specified | All participants (100% women) were injected by others, sometimes by a male partner who exerted power and control. Some participants shared needles and injecting equipment with partners as an intimate practice. Participants attributed injecting-related bacterial infections to unintentional subcutaneous injecting when self-injecting, caused by inexperience and a lack of knowledge about how to inject safely. Self-injecting was a positive experience for some women as it promoted independence; for others, it caused |
|  |  |  | participant contacts.<br>100% women; age range 16 – 46 years. |  |  | harm visible scars which worsened social marginalization. Most participants accessed a local needle exchange and had ample supply of sterile drug preparation and injecting equipment. Cleanliness and hygiene were commonly raised as important reasons to avoid reusing or sharing of equipment. |
| Small 2008<sup>66</sup> | Canada | Individual interviews<br>Thematic analysis | 50 PWID who use supervised injection sites in Vancouver.<br>42% women; median age 38; housing status not reported | Impact of the supervised consumption site on access to care for injecting-related bacterial infections. | None specified, but motivated by exploration of “social and structural barriers to care commonly experienced by [injection drug users]” | Participants described delaying or avoiding medical care because of previous negative experiences. By providing nonjudgmental care within a setting where drug use is accommodated, contact with nurses at a supervised injection site facilitated access to care for injecting-related infections. |
| Summers 2018<sup>72</sup> | USA | Mixed methods<br>Individual interviews<br>Thomas’ general inductive approach | 12 PWID recruited from needle and syringe programs in Boston, Massachusetts and Sacramento, California | Prevention and treatment of skin infections | Rhodes’ risk environment<br>Health Belief Model (HBM) of health-seeking behaviours<br>Conceptual Model of Medical Care Avoidance | Participants described delaying, avoiding, or prematurely leaving medical care for injecting-related skin and soft-tissue infections because of experiences of unaddressed pain and withdrawal symptoms, stigma. |

### Thematic synthesis

#### Summary

We identified six descriptive themes organized into two analytic themes (see Figure 3).

**Figure 3.**
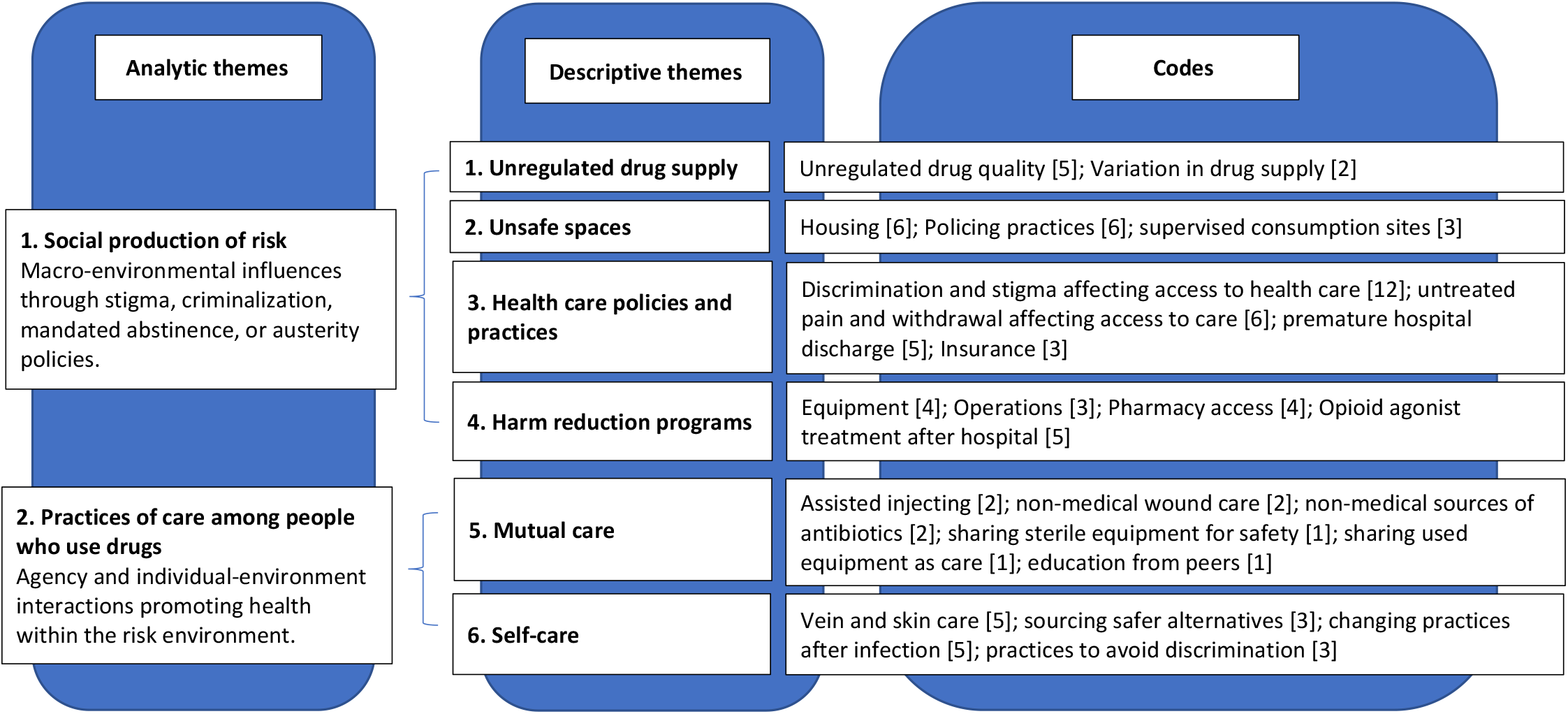
Schematic summary of analytic and descriptive themes on social and structural determinants of injection drug use-associated bacterial and fungal infections.

The first analytic theme, *social production of risk*, considers how macro-environmental factors, including criminalization, poverty, structural stigma, mandated abstinence, and racism, shape risks for injecting-related infections. Four associated descriptive themes highlighted pathways through which this occurs: (1) *unregulated drug supply*, leading to poor drug quality and solubility; (2) *unsafe spaces*, influence by insecure housing and policing practices, and ameliorated by supervised consumption sites; (3) *health care policies and practices*, leading to experiences of discrimination and undertreated pain and withdrawal, which worsened infectious complications by alienating PWID and discouraging access to care; and (4) *harm reduction programs*, including structural barriers to effective service delivery.

The second analytic theme, *practices of care among people who use drugs*, addresses PWID expertise and agency in attempts to prevent and care for bacterial infections within constraining risk environments. Two associated descriptive themes categorized these practices as (5) *mutual care*, including sharing sterile injecting equipment, assisting others with injecting into veins (rather than intramuscularly), and treating abscesses outside of medical settings; and (6) *self-care*, including promoting vein health and sourcing safer alternatives when sterile injecting equipment was unavailable. Within constraining risk environments, some of these mutual- and self-care protective strategies for bacterial infections precipitated other health risks, including HIV transmission or arterial injury.

Descriptive themes are detailed below, supplemented by quotations from study authors and participants (indicated in italics).

#### Unregulated drug supply

In five studies,^52,54–57^ authors presented perspectives from PWID who attributed infections to the quality of unregulated drugs, including adulterants,^52,54,55,57^ poor solubility,^52,54–57^ and bacterial contamination,^54^ especially through precipitating skin abscesses and vein sclerosis. Phillips and colleagues^54^ reported that PWID in Denver (USA) commonly linked their bacterial infections to poor drug quality:

> “*I think it’s the dope because… I’ll use a clean needle every time, and it still, it just depends on what they cut it with. You know, sometimes when you’re cooking it, it’s an okay color, and then the next time you’re doing it you’ve got all this shit floating up, and it’s all burnt around the sides*.” (USA)^54^

In two studies,^55,57^ authors analysed drivers of variation in the unregulated drug supply and associated infection risks. Mars and coauthors^57^ identified that PWID in Philadelphia (USA) could purchase only tar heroin (which is less soluble than powder heroin, and is associated with greater bacterial infection risk) due to regional demarcation of supply networks. Harris and colleagues^55^ highlighted London (England) PWID accounts of changing drug quality over time which has impacted widespread overuse of citric acid, which is used to dissolve poorly soluble cutting agents or adulterants such as paracetamol and quinine.

#### Unsafe spaces

In eight studies,^53,54,56,58–62^ investigators attributed increased bacterial infection risk to suboptimal drug preparation and injecting techniques created by unsafe spaces, including when PWID lacked housing and when trying to avoid being seen by police when using outdoors.

In six of these studies,^53,56,58,60,61,63^ authors explored influences of being deprived of housing on infection risk. Lack of housing made it harder to prepare and inject drugs safely, including where there were no hygienic surfaces to prepare drugs,^53,56,61^ inadequate lighting to find veins (leading to “missed hits” and inadvertent subcutaneous injection),^53^ and no clean, running water to wash hands/skin or to dissolve drugs (leading PWID to use unhygienic water alternatives)^56,58,60,61^:

“*…there was no water actually and I had to use a bit of saliva. …It worked, I still got my hit, but I also got the worst infection of my life, I nearly died* …*Yeah, I was in hospital for nearly 3 months. Septicaemia*.” (England)^56^

In their studies of PWID with endocarditis, Bearnot and colleagues^63,64^ noted that deprived of housing interfered participants’ with care, including being ineligible for outpatient parenteral antimicrobial therapy and having no fixed address for follow-up clinic contacts.

In six studies^53,54,56,58,59,62^ authors analysed how criminalizing possession of drugs or injecting equipment (and associated police enforcement) increased risk for injecting-related infections. When lacking safer indoor places to prepare and consume drugs, participants described engaging in riskier practices to avoid being seen by police. This included preparing and injecting drugs in unhygienic abandoned buildings,^58^ and compromising injecting preparation practice when hurrying and not using a filter, not using sterile water, and/or inadvertently injecting subcutaneously:^53,56,58^

“*I don’t even use cotton* [a filter]… *boom and I usually get it done. Like that. So, if the cops raid and*… *several times the cops have pulled over, come right up to me and I’ve already injected it in my arm before they hit me*.” (USA)^53^

In their ethnography, Bourgois and colleagues^59^ observed “greater and more antagonistic police surveillance” of African American PWID than of white PWID in San Francisco (USA), leading to racist, differential seizure of sterile syringes (obtained from legal needle and syringe programs). They observed police evict homeless encampments and confiscate possessions, causing PWID to miss medical appointments.

Three studies^58,65,66^ included analyses of how supervised consumption sites create safer spaces to reduce infection risks caused by lack of housing and criminalization, by facilitating individualized education on safer injecting techniques^65^ and access to wound/abscess care.^65,66^

#### Health care policies and practices

In 13 studies, authors analysed why PWID delay or avoid medical care for injecting-related infections (often until infections had progressed and spread). Contributing factors were prior experiences of stigmatizing or discriminatory care (in 12 studies^53,54,60,61,64,67–73^) and of untreated pain and withdrawal (in six studies^53,61,63,64,71,72^). In several studies, PWID described both:

> “*I’m not trying to get drugs. I’m trying to get you to take your sharp scalpel, cut this fucking thing open, squeeze this shit out of me, and get me the fuck out of here. That’s the pain relief that I want you to give me*…*I can do heroin; your little 5mg Percocet ain’t doing nothing for me. But they automatically think when you come in, ‘I got an abscess. I’m hurting’, ‘Oh, you’re trying to get drugs’, this and that… it does prevent a lot of people from going*.” (USA)^53^

Some negative experiences were driven by hospital policies. Harris^61^ explains how a London hospital policy mandates that urine drug screens be obtained before methadone can be dispensed, even if the dose is confirmed by pharmacies or treatment programs. This caused delays or missed dosages of methadone, and resulting experiences of opioid withdrawal led PWID to stay away:

> “*Mainly because how I have been treated at the hospitals, which is just like fucking dirt you’d find on your shoe… also being scared that I was going to be rough [sick]* …*because if they didn’t [give] me Methadone, like someone’s said he [doctor] won’t do it unless he would have to, and if you don’t know your rights, but yeah, it was that that really scared me more than anything, was being sick [in withdrawal] in a hospital*.” (England)^61^

Four studies^60,61,67,69^ included analyses of how delays in care due to negative experiences in health care settings had a disproportionate impact by race or gender. Assessing hospital care experiences in Vancouver (Canada), McNeil and colleagues^67^ described, “Many participants of Aboriginal ancestry further expressed that institutionalized racism reinforced the view among hospital staff that they were ‘drug-seeking’”. Three studies included descriptions of how PWID who were mothers were discouraged from accessing care for injecting-related infections, including feelings of shame at disclosing substance use as a mother,^60^ and fear of child apprehension if their substance use was reported by health professionals.^61,69^

In four studies,^63,67,70,74^ PWID described leaving hospital prematurely, before completing treatment for injecting-related infections. Explanations included leaving hospital in response to discrimination^67,74^ and because restrictions on their movements in hospital triggered post-traumatic stress.^63^ Two studies^67,70^ highlighted participants being involuntarily discharged from hospital because of drug use, despite ongoing medical need. Jafari and colleagues^74^ evaluated experiences with a care model intending to overcome these issues: clients at a residential, harm-reduction oriented program for people with severe injecting-related infections in Vancouver, Canada, described receiving less judgmental and stigmatizing care compared to their experience in mainstream hospitals.

Only one study specifically explored insufficient health insurance as a barrier to care.^69^ In other studies, authors explained that insurance is a barrier to health care for some PWID but their study participants had access to public health insurance (universally in Canada^67^, and Medicaid in USA^63,64^).

#### Harm reduction programs

In four studies,^52,55,56,71^ authors analysed consequences of PWID having insufficient or nonpreferred drug preparation and injecting equipment distributed from harm reduction programs. In their study of experiences of skin and soft-tissue infections in Glasgow (Scotland), Dunleavy and colleagues^52^ report: “reasons for re-using [needles and syringes included having been] accidentally supplied with the wrong sized needles and preferring to re-use than use the wrong needle”. Three of these studies describe PWID lacking needed equipment and repurposing alcohol skin swabs distributed by harm reduction programs: to clean up blood after injecting,^52,71^ to filter visible particulate matter out of puddle water when lacking access to sterile water,^56^ and burning swabs to obtain an adequate flame to heat the drug solution when lacking a sufficient lighter.^52^ In two studies, Harris and colleagues explored how legal/regulatory and funding restrictions on harm reduction programs limited the distribution of sterile water^56^ or single-use ascorbic acid packets.^55^

In three studies, participants described structural barriers to needle and syringe programs that limited effectiveness. This included limited operating hours (e.g. closures on weekends^52^) and restricted eligibility.^67,73^ McNeil and colleagues^67^ assessed consequences of PWID being unable to access sterile equipment in hospital, leading to reuse of contaminated equipment:

> “*[Nurses] don’t give rigs [syringes] to us. …I think that they should. If not, we’re reusing our rigs or we’re having to risk getting kicked out for stealing them or people’ll be sharing them. …I know one girl was using her same rig for days to the point where it was tearing and she was suffering every time she’d do her fix. She just didn’t have it in her to go and try and steal clean rigs*.*”* (Canada)^67^

Four further studies^62,68,75,76^ focused on places without local needle and syringe programs (in USA and Mexico), where PWID were also unable to purchase syringes at pharmacies due to refusal from pharmacists:

*“I think that many [pharmacists] think that by prohibiting the sale of syringes that they are going to stop the usage of drugs*…*but what they are doing is wrong, because of that we have a harder time finding syringes. We need to use drugs in order to feel well, since when we are in need of a fix we feel desperate enough that we don’t care and borrow one from a friend, since it’s a desperate feeling*…*”* (Mexico)^76^

Paquette and colleagues^68^ explored how PWID would prefer having multiple access points for sterile injecting equipment, including from both pharmacies and needle and syringe programs: “…one participant indicated that using the [needle and syringe program] could out him as a PWID and expose him to stigma from others because [needle and syringe programs] exclusively serve PWID. If PWID could consistently access syringes at a pharmacy without fear of discrimination, some might prefer this option because it offers a higher level of anonymity than [needle and syringe programs].”^68^

Two studies highlighted how suboptimal delivery of OAT after hospital discharge could increase risks for recurrent infections, including involuntary discharge from OAT because of ongoing use,^63^ waiting lists,^63,64^ and a lack of coordination:^64^

> “*So I had methadone maintenance while I was in the hospital and I did not really have anything lined up when I left* [hospital], *which, ultimately, could be one of the many reasons why I ended up re-infecting my valve and back in the hospital*.” (USA)^64^

#### Mutual care

Five studies^52,53,60,62,71^ included descriptions of PWID caring for each other to promote health and reduce risks of infections. Within constraining risk environments, some of these protective strategies for bacterial infections precipitated other health risks.

Mutual care practices include providing or receiving education from fellow PWID,^52^ sharing sterile needles or injecting equipment in settings of scarcity,^62^ and offering or receiving assistance with injecting to reduce infection risks^53,60^:

> “*I have my boyfriend. I only hit with him, always with him. I do not like to do it with strangers or people to whom I do not know so well. … My boyfriend helps me, because when I do it, it swells up*.” (USA)^60^
>
> Once infections developed, study participants described providing or receiving wound care and abscess treatment or antibiotics from peers in order to avoid negative experiences with the health care system.^53,71^

While navigating risk environments, protective strategies for bacterial infections could precipitate other health risks. For example, three studies^55,60,77^ assessed particular risks that women PWID face when relying on assisted injecting in the context of gendered power dynamics. In their study, Epele^60^ explored these trade-offs: “Abscesses and scars that are more frequent with muscle injection lead to further subordination within the hierarchies of their social networks, and deteriorate the women’s precarious strategies of income production. Although being injected by another increases the probability of HIV infection, it simultaneously prevents the visible physical damage that subjects these women to greater vulnerability.” Similarly, nonmedical abscess treatment or use of potentially inappropriate from nonmedical sources can lead to worsening infections, but PWID described employing these strategies to avoid negative experiences in health care settings.^53,71^

#### Self-care

Twelve studies^52,53,56–58,60,63–65,68,71,76^ included analyses of participants’ practices to prevent and treat bacterial infections. These included practices to promote vein and skin care, including staying hydrated,^71^ rotating injecting sites,^60^ taking extra time to access veins,^53,57,71^ asking for help to access veins,^60,77^ and self-treating superficial abscesses (e.g., incision and drainage; nonmedical sources of antibiotics) before they progressed.^53,68,71^

*“Little things like drink a lot of liquids, make sure you sleep every night. Make sure you get enough sleep, drink liquids, eat regularly*.*”* (USA)^71^

In three of these studies, authors highlighted actions to mitigate the risks of poor-quality drugs or injecting equipment, including sharpening the tips of used needle tips to avoid vein damage (when unable to access new needles),^76^ sourcing safer water by asking passers-by for bottled water,^56^ and using ascorbic acid (which is safer than citric acid or lemon juice) when preparing heroin.^55^

In five studies, participants also described changing their drug use practices after experiencing an infection, to avoid another one. This included applying new learnings on safer injecting techniques,^52,64,65^ switching from injecting to smoking,^52^ getting wounds assessed by a nurse,^52^ using the minimum required acidifier to dissolve drugs,^55^ and seeking addiction treatment to reduce or abstain from injection use.^63^Three studies included descriptions of self-care practices of PWID to avoid discrimination and structural stigma. This included injecting in central veins at hidden sites to avoid scars at more visible peripheral sites,^52,60^ and using in unhygienic abandoned buildings to avoid being seen.^58^ Some of these protective self-care strategies employed within constraining risk environments also led to other potential health risks. For example, injecting in central veins in the groin to avoid discrimination from visible scars increases risks of thrombosis and arterial injury, and likely increase risks for bacterial infections (as the groin has a higher burden of bacterial colonisation). Considering unintended harms of inappropriate self-treatment of bacterial infections, Gilbert and colleagues^71^ write: “There are certainly risks conferred by the self-care practices that PWID are forced to resort to. However, these risks are not taken lightly by PWID; they are weighed against the risk of inaction and worsening infections, which is well known in these communities.”

## DISCUSSION

We reviewed qualitative studies on experiences of injection drug use-associated bacterial and fungal infections, and used thematic synthesis to identify social-structural factors influencing risk for these infections. These include poor quality of unregulated drugs, insufficient housing, policing practices, limited harm reduction services, and harmful health care practices. These are shaped by macro-environmental factors including structural stigma, criminalization, government austerity, and racism. We also identified ways in which PWID care for themselves and others to prevent and treat injecting-related infections, including by sharing sterile equipment and treating infections outside of medical settings. Within constraining risk environments, PWID face trade-offs and some of these protective strategies precipitated other health risks (e.g. in some circumstances of assisted-injecting). Enabling environment interventions that address these social-structural factors could further empower people who inject drugs to protect themselves and their community. While the importance of education on safer injecting technique came up in several studies,^52,53,55,66,76^ our findings suggest that individual-level behavioural interventions alone are likely insufficient to reduce risk.

Several social-structural determinants of bacterial and fungal infections (as well as practices of mutual- and self-care^35,78^) that we identified are consistent with prior studies examining risk for HIV and HCV among PWID.^27,29,42,79^ Insecure housing, hurrying injections to avoid police, insufficient harm reduction services, and laws restricting sterile injecting equipment are known to contribute to HCV^79^ and HIV^27^ risks. Stigmatizing and discriminatory health care experiences similarly discourage HCV and HIV treatment access and exacerbate health inequities.^30,80^ Compared to the literature on HIV and HCV among PWID,^81,82^ we identified relatively little published research considering intersectionality and risk for injecting-related bacterial or fungal infections.^42^ A qualitative study by Hrycko and colleagues published after we conducted our search identified several social-structural factors contributing to risk for severe bacterial infections, including availability and use of drugs (e.g. fentanyl, stimulants) associated with a shorter duration of effect and more frequent injecting, and lack of access to sterile water.^83^

A key motivation for our review was to identify potential opportunities to reduce risks for injecting-related bacterial and fungal infections. Many social-structural factors that we identified are modifiable, and some have already been resolved or ameliorated in some places. These include PWID organizing to access better quality, regulated drugs including via injectable OAT (with liquid formulations of diacetylmorphine, hydromorphone, or fentanyl), and through “safe supply” prescribing programs or compassion clubs.^84–86^ Injectable OAT is associated with low risk for bacterial infections even when injected intramuscularly, since sterile, liquid formulations of drugs are provided in a hygienic and safe environment.^87^ Social and supportive housing (including Housing First) can help PWID access and maintain housing; some models combine housing with injectable OAT, safe supply, and/or supervised consumption sites.^88–90^ In some jurisdictions PWID and their allies have successfully advocated for decriminalization of drug/syringe possession and for laws enabling supervised consumption sites.^91^

Several initiatives have improved health care experiences for PWID with injecting-related infections,^92,93^ including incorporating harm reduction and cultural safety principles, ^61,94^ specialized addiction medicine consultation services,^95–97^ needle and syringe programs,^98,99^ and supervised consumption sites^100,101^ into hospital care. Policy changes are needed at many hospitals to facilitate these initiatives.^102,103^

Our study has three key limitations. First, our review only included studies describing experiences of injecting-related infections and we did not include all studies investigating determinants of risky injecting practices (e.g. subcutaneous injecting; reuse of contaminated equipment) unless explicitly connected to infections. Second, we did not include gray literature that might have discussed further social-structural factors beyond those we identified in peer-reviewed papers. Third, some commentators^48,104^ have argued that qualitative evidence syntheses decontextualize the nuanced findings of qualitative studies (conducted in different settings, with different methods) and try to consolidate knowledge that is not generalizable. We undertook this approach to understand how social and structural factors shape risks for injecting-related infections in ways that may be impossible to assess with quantitative research.^38,105^

## Conclusions

Injecting-related bacterial and fungal infections are shaped by modifiable social-structural factors, including unregulated drug quality, criminalization, insufficient housing, limited harm reduction services, and harmful health care practices. Enabling environment interventions that address these factors could further empower people who inject drugs to protect themselves and their community.

## Data Availability

All data produced in the present work are contained in the manuscript

## ACKNOWLEDGMENTS

We thank Louise Gillis (Research Data Librarian at Dalhousie University) for helpful feedback and assistance with our search strategy. We thank Drs. Lindsay Wallace (Cambridge University), Ashish Thakrar (University of Pennsylvania), and Paul Christine (Boston Medical Center) for helpful comments on earlier manuscript drafts.

We acknowledge that TDB, MB, EC, and IK live and work in Mi’kma’ki, the ancestral and unceded territory of the Mi’kmaq, and DW lives and works in unsurrendered and unceded territory and traditional lands of Wolastoqiyik. This territory is covered by the Treaties of Peace and Friendship which the Mi’kmaq and Wolastoqiyik Peoples first signed with the British Crown in 1725. The treaties did not deal with surrender of lands and resources but in fact recognized Mi’kmaq and Wolastoqiyik title and established the rules for what was to be an ongoing relationship between nations. We are Treaty people.

## REFERENCES

1. Brothers TD, Lewer D, Bonn M, Webster D, Harris M. Social and structural determinants of injecting-related bacterial and fungal infections among people who inject drugs: protocol for a mixed studies systematic review. BMJ Open. 2021 Aug 9;11(8):e049924. http://doi.org/10.1136/bmjopen-2021-049924

2. Gomes T, Kitchen SA, Tailor L, Men S, Murray R, Bayoumi AM, et al. Trends in Hospitalizations for Serious Infections Among People With Opioid Use Disorder in Ontario, Canada. J Addict Med. 2021 Oct 28;Publish Ahead of Print. https://doi.org/10.1097/adm.0000000000000928

3. Serota DP, Bartholomew TS, Tookes HE. Evaluating Differences in Opioid and Stimulant Use-associated Infectious Disease Hospitalizations in Florida, 2016–2017. Clin Infect Dis. 2021 Oct 1;73(7):e1649–57. https://doi.org/10.1093/cid/ciaa1278

4. Meisner JA, Anesi J, Chen X, Grande D. Changes in Infective Endocarditis Admissions in Pennsylvania During the Opioid Epidemic. Clin Infect Dis. 2020;71(7):1664–70. https://doi.org/10.1093/cid/ciz1038

5. Lewer D, Freer J, King E, Larney S, Degenhardt L, Tweed EJ, et al. Frequency of healthcare utilisation by adults who use illicit drugs: a systematic review and meta-analysis. Addiction. 2020 Jun;115(6):1011–23. https://doi.org/10.1111/add.14892

6. Kim JH, Fine DR, Li L, Kimmel SD, Ngo LH, Suzuki J, et al. Disparities in United States hospitalizations for serious infections in patients with and without opioid use disorder: A nationwide observational study. PLoS Med. 2020 Aug 7;17(8):e1003247. https://doi.org/10.1371/journal.pmed.1003247

7. Wright A, Otome O, Harvey C, Bowe S, Athan E. The Current Epidemiology of Injecting Drug Use-Associated Infective Endocarditis in Victoria, Australia in the Midst of Increasing Crystal Methamphetamine Use. Heart Lung Circ. 2018 Apr 1;27(4):484–8. https://doi.org/10.1016/j.hlc.2017.03.166

8. Mosseler K, Materniak S, Brothers TD, Webster D. Epidemiology, microbiology, and clinical outcomes among patients with intravenous drug use-associated infective endocarditis in New Brunswick. Can J Cardiol Open. 2020;2(5):379–85. https://doi.org/10.1016/j.cjco.2020.05.002

9. Lewer D, Harris M, Hope V. Opiate Injection–Associated Skin, Soft Tissue, and Vascular Infections, England, UK, 1997–2016. Emerg Infect Dis. 2019 Sep 23;23(8):1400–3. https://doi.org/10.3201/eid2308.170439

10. Wurcel AG, Anderson JE, Chui KKH, Skinner S, Knox TA, Snydman DR, et al. Increasing Infectious Endocarditis Admissions Among Young People Who Inject Drugs. Open Forum Infect Dis. 2016 Jul 26;3(3):ofw157. https://doi.org/10.1093/ofid/ofw157

11. Barocas JA, Eftekhari Yazdi G, Savinkina A, Nolen S, Savitzky C, Samet JH, et al. Long-term Infective Endocarditis Mortality Associated With Injection Opioid Use in the United States: A Modeling Study. Clin Infect Dis. 2021 Dec 6;73(11):ciaa1346. https://doi.org/10.1093/cid/ciaa1346

12. McCarthy NL, Baggs J, See I, Reddy SC, Jernigan JA, Gokhale RH, et al. Bacterial Infections Associated With Substance Use Disorders, Large Cohort of United States Hospitals, 2012– 2017. Clin Infect Dis. 2020 Oct 23;71(7):e37–44. https://doi.org/10.1093/cid/ciaa008

13. Cooper HLF, Brady JE, Ciccarone D, Tempalski B, Gostnell K, Friedman SR. Nationwide Increase in the Number of Hospitalizations for Illicit Injection Drug Use-Related Infective Endocarditis. Clin Infect Dis. 2007 Nov 1;45(9):1200–3. https://doi.org/10.1086/522176

14. Ronan MV, Herzig SJ. Hospitalizations Related To Opioid Abuse/Dependence And Associated Serious Infections Increased Sharply, 2002–12. Health Aff (Millwood). 2016 Jan 5;35(5):832–7. https://doi.org/10.1377/hlthaff.2015.1424

15. Larney S, Peacock A, Mathers BM, Hickman M, Degenhardt L. A systematic review of injecting-related injury and disease among people who inject drugs. Drug Alcohol Depend. 2017 Feb 1;171:39–49. https://doi.org/10.1016/j.drugalcdep.2016.11.029

16. Gordon RJ, Lowy FD. Bacterial Infections in Drug Users. N Engl J Med. 2005 Nov 3;353(18):1945–54. https://doi.org/10.1056/NEJMra042823

17. Dwyer R, Topp L, Maher L, Power R, Hellard M, Walsh N, et al. Prevalences and correlates of non-viral injecting-related injuries and diseases in a convenience sample of Australian injecting drug users. Drug Alcohol Depend. 2009 Feb 1;100(1):9–16. https://doi.org/10.1016/j.drugalcdep.2008.08.016

18. Hope VD, Hickman M, Parry JV, Ncube F. Factors associated with recent symptoms of an injection site infection or injury among people who inject drugs in three English cities. Int J Drug Policy. 2014 Mar 1;25(2):303–7. https://doi.org/10.1016/j.drugpo.2013.11.012

19. Phillips KT, Stein MD. Risk Practices Associated with Bacterial Infections among Injection Drug Users in Denver, Colorado. Am J Drug Alcohol Abuse. 2010 Mar 1;36(2):92–7. https://doi.org/10.3109/00952991003592311

20. Roux P, Donadille C, Magen C, Schatz E, Stranz R, Curano A, et al. Implementation and evaluation of an educational intervention for safer injection in people who inject drugs in Europe: a multi-country mixed-methods study. Int J Drug Policy. 2021 Jan 1;87:102992. https://doi.org/10.1016/j.drugpo.2020.102992

21. Stein MD, Phillips KT, Herman DS, Keosaian J, Stewart C, Anderson BJ, et al. Skin-cleaning among hospitalized people who inject drugs: a randomized controlled trial. Addiction. 2021 May;116(5):122–1130. https://doi.org/10.1111/add.15236

22. Phillips KT, Stewart C, Anderson BJ, Liebschutz JM, Herman DS, Stein MD. A randomized controlled trial of a brief behavioral intervention to reduce skin and soft tissue infections among people who inject drugs. Drug Alcohol Depend. 2021 Feb 27;108646. https://doi.org/10.1016/j.drugalcdep.2021.108646

23. Harvey M. The Political Economy of Health: Revisiting Its Marxian Origins to Address 21st-Century Health Inequalities. Am J Public Health. 2021 Feb;111(2):293–300. https://ajph.aphapublications.org/doi/full/10.2105/AJPH.2020.305996

24. Holman D, Lynch R, Reeves A. How do health behaviour interventions take account of social context? A literature trend and co-citation analysis. Health (N Y). 2018 Jul 1;22(4):389–410. https://doi.org/10.1177/1363459317695630

25. Link BG, Phelan J. Social Conditions As Fundamental Causes of Disease. J Health Soc Behav. 1995;35:80. https://doi.org/10.2307/2626958

26. Krieger N. Theories for social epidemiology in the 21st century: an ecosocial perspective. Int J Epidemiol. 2001 Aug 1;30(4):668–77. https://doi.org/10.1093/ije/30.4.668

27. Strathdee SA, Hallett TB, Bobrova N, Rhodes T, Booth R, Abdool R, et al. HIV and risk environment for injecting drug users: the past, present, and future. Lancet Lond Engl. 2010 Jul 24;376(9737):268–84. https://doi.org/10.1016/s0140-6736(10)60743-x

28. Rhodes T, Simic M. Transition and the HIV risk environment. BMJ. 2005 Jul 23;331(7510):220–3. https://doi.org/10.1136/bmj.331.7510.220

29. Rhodes T, Singer M, Bourgois P, Friedman SR, Strathdee SA. The social structural production of HIV risk among injecting drug users. Soc Sci Med. 2005 Sep 1;61(5):1026–44. https://doi.org/10.1016/j.socscimed.2004.12.024

30. Harris M, Rhodes T. Hepatitis C treatment access and uptake for people who inject drugs: a review mapping the role of social factors. Harm Reduct J. 2013 May 7;10(1):7. https://doi.org/10.1186/1477-7517-10-7

31. Dasgupta N, Beletsky L, Ciccarone D. Opioid Crisis: No Easy Fix to Its Social and Economic Determinants. Am J Public Health. 2017 Dec 21;108(2):182–6. https://doi.org/10.2105/ajph.2017.304187

32. McLean K. “There’s nothing here”: Deindustrialization as risk environment for overdose. Int J Drug Policy. 2016 Mar 1;29:19–26. https://doi.org/10.1016/j.drugpo.2016.01.009

33. Amemiya J, Mortenson E, Heyman GD, Walker CM. Thinking Structurally: A Cognitive Framework for Understanding How People Attribute Inequality to Structural Causes. Perspect Psychol Sci. 2022 Aug 18;17456916221093592. https://doi.org/10.1177/17456916221093593

34. McNeil R, Small W. ‘Safer environment interventions’: A qualitative synthesis of the experiences and perceptions of people who inject drugs. Soc Sci Med. 2014 Apr;106:151–8. https://doi.org/10.1016/j.socscimed.2014.01.051

35. Harris M, Rhodes T. Venous access and care: harnessing pragmatics in harm reduction for people who inject drugs. Addiction. 2012 Jun;107(6):1090–6. https://doi.org/10.1111/j.1360-0443.2011.03749.x

36. Page MJ, McKenzie JE, Bossuyt PM, Boutron I, Hoffmann TC, Mulrow CD, et al. The PRISMA 2020 statement: an updated guideline for reporting systematic reviews. BMJ. 2021 Mar 29;372:n71. https://doi.org/10.1136/bmj.n71

37. Tong A, Flemming K, McInnes E, Oliver S, Craig J. Enhancing transparency in reporting the synthesis of qualitative research: ENTREQ. BMC Med Res Methodol. 2012 Nov 27;12(1):181. https://doi.org/10.1186/1471-2288-12-181

38. Pluye P, Hong QN. Combining the Power of Stories and the Power of Numbers: Mixed Methods Research and Mixed Studies Reviews. Annu Rev Public Health. 2014;35(1):29–45. https://doi.org/10.1146/annurev-publhealth-032013-182440

39. Pluye P, Hong QN, Bush PL, Vedel I. Opening-up the definition of systematic literature review: the plurality of worldviews, methodologies and methods for reviews and syntheses. J Clin Epidemiol. 2016 May 1;73:2–5. https://doi.org/10.1016/j.jclinepi.2015.08.033

40. Rhodes T. The ‘risk environment’: a framework for understanding and reducing drug-related harm. Int J Drug Policy. 2002 Jun 1;13(2):85–94. https://doi.org/10.1016/S0955-3959(02)00007-5

41. Rhodes T. Risk environments and drug harms: A social science for harm reduction approach. Int J Drug Policy. 2009 May 1;20(3):193–201. https://doi.org/10.1016/j.drugpo.2008.10.003

42. Collins AB, Boyd J, Cooper HLF, McNeil R. The intersectional risk environment of people who use drugs. Soc Sci Med. 2019 Aug 1;234:112384. https://doi.org/10.1016/j.socscimed.2019.112384

43. Rhodes T, Wagner K, Strathdee SA, Shannon K, Davidson P, Bourgois P. Structural Violence and Structural Vulnerability Within the Risk Environment: Theoretical and Methodological Perspectives for a Social Epidemiology of HIV Risk Among Injection Drug Users and Sex Workers. In: O’Campo P, Dunn JR, editors. Rethinking Social Epidemiology: Towards a Science of Change. Dordrecht: Springer Netherlands; 2012. p. 205–30. https://doi.org/10.1007/978-94-007-2138-8_10

44. Robertson R, Broers B, Harris M. Injecting drug use, the skin and vasculature. Addiction. 2021 Jul;116(7):add.15283. https://doi.org/10.1111/add.15283

45. Moola S, Munn Z, Sears K, Sfetcu R, Currie M, Lisy K, et al. Conducting systematic reviews of association (etiology): The Joanna Briggs Institute’s approach. JBI Evid Implement. 2015 Sep;13(3):163–9. https://doi.org/10.1097/xeb.0000000000000064

46. Hong QN, Fàbregues S, Bartlett G, Boardman F, Cargo M, Dagenais P, et al. The Mixed Methods Appraisal Tool (MMAT) version 2018 for information professionals and researchers. Educ Inf. 2018 Jan 1;34(4):285–91. https://content.iospress.com/articles/education-for-information/efi180221

47. Hong QN, Pluye P. A Conceptual Framework for Critical Appraisal in Systematic Mixed Studies Reviews. J Mix Methods Res. 2019 Oct 1;13(4):446–60. https://doi.org/10.1177/1558689818770058

48. Thomas J, Harden A. Methods for the thematic synthesis of qualitative research in systematic reviews. BMC Med Res Methodol. 2008 Jul 10;8(1):45. https://doi.org/10.1186/1471-2288-8-45

49. Guise A, Horyniak D, Melo J, McNeil R, Werb D. The experience of initiating injection drug use and its social context: a qualitative systematic review and thematic synthesis. Addiction. 2017;112(12):2098–111. https://doi.org/10.1111/add.13957

50. Harris M, Guy D, Picchio CA, White TM, Rhodes T, Lazarus JV. Conceptualising hepatitis C stigma: A thematic synthesis of qualitative research. Int J Drug Policy. 2021 Oct 1;96:103320. https://doi.org/10.1016/j.drugpo.2021.103320

51. Yoon GH, Levengood TW, Davoust MJ, Ogden SN, Kral AH, Cahill SR, et al. Implementation and sustainability of safe consumption sites: a qualitative systematic review and thematic synthesis. Harm Reduct J. 2022 Jul 5;19(1):73. https://doi.org/10.1186/s12954-022-00655-z

52. Dunleavy K, Hope V, Roy K, Taylor A. The experiences of people who inject drugs of skin and soft tissue infections and harm reduction: A qualitative study. Int J Drug Policy. 2019;65:65–72. https://doi.org/10.1016/j.drugpo.2018.09.001

53. Harris RE, Richardson J, Frasso R, Anderson ED. Experiences with skin and soft tissue infections among people who inject drugs in Philadelphia: A qualitative study. Drug Alcohol Depend. 2018 Jun 1;187:8–12. https://doi.org/10.1016/j.drugalcdep.2018.01.029

54. Phillips KT, Altman JK, Corsi KF, Stein MD. Development of a risk reduction intervention to reduce bacterial and viral infections for injection drug users. Subst Use Misuse. 2013 Jan;48(1–2):54–64.

55. Harris M, Scott J, Wright T, Brathwaite R, Ciccarone D, Hope V. Injecting-related health harms and overuse of acidifiers among people who inject heroin and crack cocaine in London: a mixed-methods study. Harm Reduct J. 2019 Nov 13;16(1):60. https://doi.org/10.1186/s12954-019-0330-6

56. Harris M, Scott J, Hope V, Wright T, McGowan C, Ciccarone D. Navigating environmental constraints to injection preparation: the use of saliva and other alternatives to sterile water among unstably housed PWID in London. Harm Reduct J. 2020 Apr 10;17(1):24. https://doi.org/10.1186/s12954-020-00369-0

57. Mars SG, Bourgois P, Karandinos G, Montero F, Ciccarone D. The Textures of Heroin: User Perspectives on “Black Tar” and Powder Heroin in Two U.S. Cities. J Psychoact Drugs. 2016;48(4):270–8. https://doi.org/10.1080/02791072.2016.1207826

58. Harris RE, Richardson J, Frasso R, Anderson ED. Perceptions about supervised injection facilities among people who inject drugs in Philadelphia. Int J Drug Policy. 2018;52:56–61.

59. Bourgois P, Martinez A, Kral A, Edlin BR, Schonberg J, Ciccarone D. Reinterpreting ethnic patterns among white and African American men who inject heroin: a social science of medicine approach. PLoS Med. 2006 Oct;3(10):e452.

60. Epele ME. Scars, harm and pain: About being injected among drug using Latina women. J Ethn Subst Abuse. 2002;1(1):47–69. https://doi.org/10.1300/J233v01n01_04

61. Harris M. Normalised pain and severe health care delay among people who inject drugs in London: Adapting cultural safety principles to promote care. Soc Sci Med. 2020 Sep 1;260:113183. https://doi.org/10.1016/j.socscimed.2020.113183

62. Pollini RA, Paquette CE, Slocum S, LeMire D. ‘It’s just basically a box full of disease’-navigating sterile syringe scarcity in a rural New England state. Addict Abingdon Engl. 2021 Jan;116(1):107–15. https://doi.org/10.1111/add.15113

63. Bearnot B, Mitton JA. “You’re Always Jumping Through Hoops”: Journey Mapping the Care Experiences of Individuals With Opioid Use Disorder-associated Endocarditis. J Addict Med. 2020 Apr 7;Publish Ahead of Print. https://doi.org/10.1097/adm.0000000000000648

64. Bearnot B, Mitton JA, Hayden M, Park ER. Experiences of care among individuals with opioid use disorder-associated endocarditis and their healthcare providers: Results from a qualitative study. J Subst Abuse Treat. 2019 Jul 1;102:16–22. https://doi.org/10.1016/j.jsat.2019.04.008

65. Krüsi A, Small W, Wood E, Kerr T. An integrated supervised injecting program within a care facility for HIV-positive individuals: a qualitative evaluation. AIDS Care. 2009 May;21(5):638–44. https://doi.org/10.1080/09540120802385645

66. Small W, Wood E, Lloyd-Smith E, Tyndall M, Kerr T. Accessing care for injection-related infections through a medically supervised injecting facility: A qualitative study. Drug Alcohol Depend. 2008 Nov 1;98(1):159–62. https://doi.org/10.1016/j.drugalcdep.2008.05.014

67. McNeil R, Small W, Wood E, Kerr T. Hospitals as a “risk environment”: An ethno-epidemiological study of voluntary and involuntary discharge from hospital against medical advice among people who inject drugs. Soc Sci Med. 2014 Mar;105:59–66. https://doi.org/10.1016/j.socscimed.2014.01.010

68. Paquette CE, Syvertsen JL, Pollini RA. Stigma at every turn: Health services experiences among people who inject drugs. Int J Drug Policy. 2018 Jul;57:104–10. https://doi.org/10.1016/j.drugpo.2018.04.004

69. Colwill JP, Sherman MI, Siedlecki SL, Burchill CN, Siegmund LA. A grounded theory approach to the care experience of patients with intravenous drug use/abuse-related endocarditis. Appl Nurs Res. 2021;57:N.PAG-N.PAG. https://doi.org/10.1016/j.apnr.2020.151390

70. Bodkin K, Delahunty-Pike A, O’Shea T. Reducing stigma in healthcare and law enforcement: A novel approach to service provision for street level sex workers. Int J Equity Health. 2015;14(1). https://doi.org/10.1186/s12939-015-0156-0

71. Gilbert AR, Hellman JL, Wilkes MS, Rees VW, Summers PJ. Self-care habits among people who inject drugs with skin and soft tissue infections: a qualitative analysis. Harm Reduct J. 2019 Dec 12;16(1):69. https://doi.org/10.1186/s12954-019-0345-z

72. Summers PJ, Hellman JL, MacLean MR, Rees VW, Wilkes MS. Negative experiences of pain and withdrawal create barriers to abscess care for people who inject heroin. A mixed methods analysis. Drug Alcohol Depend. 2018 Sep 1;190:200–8. https://doi.org/10.1016/j.drugalcdep.2018.06.010

73. Meyer JP, Culbert GJ, Azbel L, Bachireddy C, Kurmanalieva A, Rhodes T, et al. A qualitative study of diphenhydramine injection in Kyrgyz prisons and implications for harm reduction. Harm Reduct J. 2020;17(1):86. https://doi.org/10.1186/s12954-020-00435-7

74. Jafari S, Joe R, Elliot D, Nagji A, Hayden S, Marsh DC. A Community Care Model of Intravenous Antibiotic Therapy for Injection Drug Users with Deep Tissue Infection for “Reduce Leaving Against Medical Advice.” Int J Ment Health Addict. 2015;13:49–58. https://doi.org/10.1007/s11469-014-9511-4

75. Case P, Ramos R, Brouwer KC, Firestone-Cruz M, Pollini RA, Fraga MA, et al. At the borders, on the edge: use of injected methamphetamine in Tijuana and Ciudad Juarez, Mexico. J Immigr Minor Health. 2008 Feb;10(1):23–33. https://doi.org/10.1007/s10903-007-9051-0

76. Pollini RA, Lozada R, Gallardo M, Rosen P, Vera A, Macias A, et al. Barriers to pharmacy-based syringe purchase among injection drug users in Tijuana, Mexico: a mixed methods study. AIDS Behav. 2010 Jun;14(3):679–87. https://doi.org/10.1007/s10461-010-9674-3

77. Sheard L, Tompkins C. Contradictions and Misperceptions: An Exploration of Injecting Practice, Cleanliness, Risk, and Partnership in the Lives of Women Drug Users. Qual Health Res. 2008 Nov 1;18(11):1536–47. https://doi.org/10.1177/1049732308325838

78. Kolla G, Strike C. Practices of care among people who buy, use, and sell drugs in community settings. Harm Reduct J. 2020 May 7;17(1):27. https://doi.org/10.1186/s12954-020-00372-5

79. Rhodes T, Treloar C. The social production of hepatitis C risk among injecting drug users: a qualitative synthesis. Addiction. 2008 Oct;103(10):1593–603. https://doi.org/10.1111/j.1360-0443.2008.02306.x

80. Krüsi A, Wood E, Montaner J, Kerr T. Social and structural determinants of HAART access and adherence among injection drug users. Int J Drug Policy. 2010 Jan 1;21(1):4–9. https://doi.org/10.1016/j.drugpo.2009.08.003

81. Bluthenthal RN. Structural racism and violence as social determinants of health: Conceptual, methodological and intervention challenges. Drug Alcohol Depend. 2021 May;222:108681. https://doi.org/10.1016/j.drugalcdep.2021.108681

82. Touesnard N, Brothers TD, Bonn M, Edelman EJ. Overdose deaths and HIV infections among people who use drugs: shared determinants and integrated responses. Expert Rev Anti Infect Ther. 2022 May 26;0(0):1–5. https://doi.org/10.1080/14787210.2022.2081152

83. Hrycko A, Mateu-Gelabert P, Ciervo C, Linn-Walton R, Eckhardt B. Severe bacterial infections in people who inject drugs: the role of injection-related tissue damage. Harm Reduct J. 2022 May 2;19(1):41. https://doi.org/10.1186/s12954-022-00624-6

84. Bonn M, Palayew A, Bartlett S, Brothers TD, Touesnard N, Tyndall M. Addressing the Syndemic of HIV, Hepatitis C, Overdose, and COVID-19 Among People Who Use Drugs: The Potential Roles for Decriminalization and Safe Supply. J Stud Alcohol Drugs. 2020 Sep 1;81(5):556–60. https://doi.org/10.15288/jsad.2020.81.556

85. BCCSU. Heroin Compassion Clubs: A cooperative model to reduce opioid overdose deaths & disrupt organized crime’s role in fentanyl, money laundering & housing unaffordability [Internet]. Vancouver, B.C.: British Columbia Centre on Substance Use.; https://www.bccsu.ca/wp-content/uploads/2019/02/Report-Heroin-Compassion-Clubs.pdf

86. Canadian Association of People Who Use Drugs (CAPUD). Safe Supply Concept Document [Internet]. 2019 Feb. https://doi.org/10.5281/zenodo.5637607

87. Meyer M, Eichenberger R, Strasser J, Dürsteler KM, Vogel M. «One prick and then it’s done»: a mixed-methods exploratory study on intramuscular injection in heroin-assisted treatment. Harm Reduct J. 2021 Dec 18;18(1):134. https://doi.org/10.1186/s12954-021-00584-3

88. Harris MT, Seliga RK, Fairbairn N, Nolan S, Walley AY, Weinstein ZM, et al. Outcomes of Ottawa, Canada’s Managed Opioid Program (MOP) where supervised injectable hydromorphone was paired with assisted housing. Int J Drug Policy. 2021 Dec 1;98:103400. https://doi.org/10.1016/j.drugpo.2021.103400

89. Brothers TD, Leaman M, Bonn M, John Fraser, Amy Gillis, Michael Gniewek, et al. Evaluation of an emergency safe supply drugs and managed alcohol program in COVID-19 isolation hotel shelters for people experiencing homelessness. Drug Alcohol Depend. 2022 Jun 1;235:109440. https://doi.org/10.1016/j.drugalcdep.2022.109440

90. Bardwell G, Collins AB, McNeil R, Boyd J. Housing and overdose: an opportunity for the scale-up of overdose prevention interventions? Harm Reduct J. 2017 Dec 6;14(1):77. https://doi.org/10.1186/s12954-017-0203-9

91. McNeil R, Small W, Lampkin H, Shannon K, Kerr T. “People Knew They Could Come Here to Get Help”: An Ethnographic Study of Assisted Injection Practices at a Peer-Run ‘Unsanctioned’ Supervised Drug Consumption Room in a Canadian Setting. AIDS Behav. 2014 Mar 1;18(3):473–85. https://doi.org/10.1007/s10461-013-0540-y

92. Brothers TD, Fraser J, Webster D. Caring for people who inject drugs when they are admitted to hospital. CMAJ Can Med Assoc J. 2021 Mar 22;193(12):E423–4. https://doi.org/10.1503/cmaj.202124

93. CRISM. Guidance Document on the Management of Substance Use in Acute Care [Internet]. Alberta: Canadian Research Initiative on Substance Misuse (CRISM) - Prairie Node; 2020. https://crismprairies.ca/management-of-substance-use-in-acute-care-settings-in-alberta-guidance-document/

94. McCall J, Pauly B. Sowing a Seed of Safety: Providing Culturally Safe Care in Acute Care Settings for People who use Drugs. J Ment Health Addict Nurs. 2019 May 31;3(1):e1–7. https://doi.org/10.22374/jmhan.v3i1.33

95. Englander H, Englander H. “We’ve Learned It’s a Medical Illness, Not a Moral Choice”: Qualitative Study of the Effects of a Multicomponent Addiction Intervention on Hospital Providers’ Attitudes and Experiences. J Hosp Med. 2018 Nov 1;13(11). https://doi.org/10.12788/jhm.2993

96. Brothers T, Fraser J, MacAdam E, Hickcox S, L Genge, T O’Donnell, et al. Implementation and evaluation of a novel, unofficial, trainee-organized hospital addiction medicine consultation service. Subst Abuse. 2021;42(4):433–7. https://doi.org/10.1080/08897077.2020.1856291

97. Hyshka E, Morris H, Anderson-Baron J, Nixon L, Dong K, Salvalaggio G. Patient perspectives on a harm reduction-oriented addiction medicine consultation team implemented in a large acute care hospital. Drug Alcohol Depend. 2019 Nov 1;204:107523. https://doi.org/10.1016/j.drugalcdep.2019.06.025

98. Brooks HL, O’Brien DC, Salvalaggio G, Dong K, Hyshka E. Uptake into a bedside needle and syringe program for acute care inpatients who inject drugs. Drug Alcohol Rev. 2019;38(4):423–7. https://doi.org/10.1111/dar.12930

99. Brothers TD, Mosseler K, Kirkland S, Melanson P, Barrett L, Webster D. Unequal access to opioid agonist treatment and sterile injecting equipment among hospitalized patients with injection drug use-associated infective endocarditis. PLoS ONE. 2022 Jan 26;17(1):e0263156. https://doi.org/10.1371/journal.pone.0263156

100. Dong KA, Brouwer J, Johnston C, Hyshka E. Supervised consumption services for acute care hospital patients. CMAJ. 2020 May 4;192(18):E476–9. https://doi.org/10.1503/cmaj.191365

101. Dogherty E, Patterson C, Gagnon M, Harrison S, Chase J, Boerstler J, et al. Implementation of a nurse-led overdose prevention site in a hospital setting: lessons learned from St. Paul’s Hospital, Vancouver, Canada. Harm Reduct J. 2022 Feb 5;19(1):13. https://doi.org/10.1186/s12954-022-00596-7

102. Lennox R, Martin L, Brimner C, O’Shea T. Hospital policy as a harm reduction intervention for people who use drugs. Int J Drug Policy. 2021 Nov 1;97:103324. https://doi.org/10.1016/j.drugpo.2021.103324

103. Harris M, Holland A, Lewer D, Brown M, Eastwood N, Sutton G, et al. Barriers to management of opioid withdrawal in hospitals in England: a document analysis of hospital policies on the management of substance dependence. BMC Med. 2022 Apr 14;20(1):151. https://doi.org/10.1186/s12916-022-02351-y

104. Dixon-Woods M, Bonas S, Booth A, Jones DR, Miller T, Sutton AJ, et al. How can systematic reviews incorporate qualitative research? A critical perspective. Qual Res. 2006 Feb 1;6(1):27–44. https://doi.org/10.1177/1468794106058867

105. Bjerre-Nielsen A, Glavind KL. Ethnographic data in the age of big data: How to compare and combine. Big Data Soc. 2022 Jan 1;9(1):20539517211069892. https://doi.org/10.1177/20539517211069893

