## supplementary Table for "Social and structural determinants of injection drug use-associated bacterial and fungal infections: a qualitative systematic review and thematic synthesis"

Supplementary appendix to:

**Social and structural determinants of injecting-related bacterial and fungal infections among people who inject drugs: a qualitative systematic review and thematic synthesis**

Thomas D. Brothers^1,2^, Matthew Bonn^3^, Dan Lewer^1^, Emilie Comeau^4^, Inhwa Kim^4^, Duncan Webster^2,5^, Andrew Hayward^1^, Magdalena Harris^6^

^1^UCL Collaborative Centre for Inclusion Health, Institute of Epidemiology and Health Care, University College London, London, UK

^2^Department of Medicine, Faculty of Medicine, Dalhousie University, Halifax, Canada

^3^Canadian Association of People who Use Drugs (CAPUD), Dartmouth, Canada

^4^Faculty of Medicine, Dalhousie University, Halifax, Canada

^5^Division of Infectious Diseases, Saint John Regional Hospital, Saint John, Canada

^6^Department of Public Health, Environments and Society, London School of Hygiene & Tropical Medicine, London, UK

### Supplementary Table 1. Full search strategy.

| **Concepts** | **PubMed MEDLINE** | **EMBASE** | **Scopus** | **CINAHL** | **PsycINFO** |
| --- | --- | --- | --- | --- | --- |
| People who inject drugs, or drug preparation and injection | (“Substance-Related Disorders”[MeSH] OR | ('substance abuse'/exp OR | ( | ( |  |
|  | “Substance Abuse, Intravenous”[MeSH] OR | 'intravenous drug abuse'/exp OR |  |  |  |
|  | “Drug Users”[MeSH] OR | 'drug use'/exp OR |  |  |  |
|  | “Needle Sharing”[MeSH] OR | 'needle sharing'/exp OR |  |  |  |
|  | “people who inject drugs”[tiab] OR “persons who inject drugs”[tiab] OR PWID[tiab] OR | “people who inject drugs”:ab,ti OR “persons who inject drugs”:ab,ti OR PWID:ab,ti OR | TITLE-ABS("people who inject drugs") OR TITLE-ABS("persons who inject drugs") OR TITLE-ABS("PWID") OR | TI("people who inject drugs" OR "persons who inject drugs" OR "PWID") OR AB("people who inject drugs" OR "persons who inject drugs" OR "PWID") OR | TI("people who inject drugs" OR "persons who inject drugs" OR "PWID") OR AB("people who inject drugs" OR "persons who inject drugs" OR "PWID") OR |
|  | “people who use drugs”[tiab] OR “persons who use drugs”[tiab] OR PWUD[tiab] OR | “people who use drugs”:ab,ti OR “persons who use drugs”:ab,ti OR PWUD:ab,ti OR | TITLE-ABS("people who use drugs") OR TITLE-ABS("persons who use drugs") OR TITLE-ABS("PWUD") OR | TI("people who use drugs" OR "persons who use drugs" OR "PWUD") OR AB("people who use drugs" OR "persons who use drugs" OR "PWUD") OR | TI("people who use drugs" OR "persons who use drugs" OR "PWUD") OR AB("people who use drugs" OR "persons who use drugs" OR "PWUD") OR |
|  | “injection drug”[tiab] OR IDU[tiab] OR | “injection drug”:ab,ti OR IDU:ab,ti OR | TITLE-ABS("injection drug") OR TITLE-ABS("IDU") OR | TI("injection drug" OR "IDU") OR AB("injection drug" OR "IDU") OR | TI("injection drug" OR "IDU") OR AB("injection drug" OR "IDU") OR |
|  | “intravenous drug”[tiab] OR IVDU[tiab] OR | “intravenous drug”:ab,ti OR IVDU:ab,ti OR | TITLE-ABS("intravenous drug") OR TITLE-ABS("IVDU") OR | TI("intravenous drug" OR "IVDU") OR AB("intravenous drug" OR "IVDU") OR | TI("intravenous drug" OR "IVDU") OR AB("intravenous drug" OR "IVDU") OR |
|  | “drug abuse”[tiab] OR | “drug abuse”:ab,ti OR | TITLE-ABS("drug abuse") OR | TI("drug abuse") OR AB("drug abuse") OR | TI("drug abuse") OR AB("drug abuse") OR |
|  | “illicit drugs”[MeSH] OR “illicit drug”[tiab] OR | 'illicit drug'/exp OR “illicit drugs”:ab,ti OR | TITLE-ABS("illicit drug") OR | TI("illicit drug") OR AB("illicit drug") OR | TI("illicit drug") OR AB("illicit drug") OR |
|  | “Heroin”[MeSH] OR Heroin[tiab] OR | “heroin”:ab,ti OR | TITLE-ABS("Heroin") OR | TI("Heroin") OR AB("Heroin") OR | TI("Heroin") OR AB("Heroin") OR |
|  | “Heroin Dependence”[MeSH] OR | 'heroin dependence'/exp OR |  |  |  |
|  | “Opiate use disorder”[tiab] OR “opioid use disorder”[tiab] OR “opiate dependence”[tiab] OR “opioid dependence”[tiab] OR “opiate abuse”[tiab] OR “opioid abuse”[tiab] OR | 'narcotic dependence'/exp OR “opioid use disorder”:ab,ti OR “opiate use disorder”:ab,ti OR | TITLE-ABS("Opiate use disorder") OR TITLE-ABS("opioid use disorder") OR TITLE-ABS("opiate dependence") OR TITLE-ABS("opioid dependence") OR TITLE-ABS("opiate abuse") OR TITLE-ABS("opioid abuse") OR | TI("Opiate use disorder" OR "opioid use disorder" OR "opiate dependence" OR "opioid dependence" OR "opiate abuse" OR "opioid abuse") OR AB("Opiate use disorder" OR "opioid use disorder" OR "opiate dependence" OR "opioid dependence" OR "opiate abuse" OR "opioid abuse") OR | TI("Opiate use disorder" OR "opioid use disorder" OR "opiate dependence" OR "opioid dependence" OR "opiate abuse" OR "opioid abuse") OR AB("Opiate use disorder" OR "opioid use disorder" OR "opiate dependence" OR "opioid dependence" OR "opiate abuse" OR "opioid abuse") OR |
|  | “Cocaine”[MeSH] OR cocaine[tiab] OR | 'cocaine'/exp OR 'cocaine dependence'/exp OR cocaine:ab,ti OR | TITLE-ABS("cocaine") OR | TI("cocaine") OR AB("cocaine") OR | TI("cocaine") OR AB("cocaine") OR |
|  | “Crack Cocaine”[MeSH] OR | “crack cocaine”:ab,ti OR | TITLE-ABS("crack cocaine") OR | TI("crack cocaine") OR AB("crack cocaine") OR | TI("crack cocaine") OR AB("crack cocaine") OR |
|  | “groin injecting”[tiab] OR “femoral injecting”[tiab] OR | “groin injecting”:ab,ti OR “femoral injecting”:ab,ti OR | TITLE-ABS("groin injecting") OR TITLE-ABS("femoral injecting") OR | TI("groin injecting" OR "femoral injecting") OR AB("groin injecting" OR "femoral injecting") OR | TI("groin injecting" OR "femoral injecting") OR AB("groin injecting" OR "femoral injecting") OR |
|  | “Harm Reduction”[MeSH] OR “harm reduction”[tiab] OR | 'harm reduction'/exp OR “harm reduction”:ab,ti OR | TITLE-ABS("harm reduction") OR | TI("harm reduction") OR AB("harm reduction") OR | TI("harm reduction") OR AB("harm reduction") OR |
|  | “Needle-Exchange Programs”[MeSH] OR |  |  |  |  |
|  | “needle exchange”[tiab] OR “syringe exchange”[tiab] OR “syringe services”[tiab] OR | “needle exchange”:ab,ti OR “syringe exchange”:ab,ti OR “syringe services”:ab,ti OR | TITLE-ABS("needle exchange") OR TITLE-ABS("syringe exchange") OR TITLE-ABS("syringe services") OR | TI("needle exchange" OR "syringe exchange") OR "syringe services") OR AB("needle exchange" OR "syringe exchange") OR "syringe services") OR | TI("needle exchange" OR "syringe exchange") OR "syringe services") OR AB("needle exchange" OR "syringe exchange") OR "syringe services") OR |
|  | acidifier*[tiab] OR | acidifier*:ab,ti OR | TITLE-ABS("acidifier*") OR | TI("acidifier*") OR AB("acidifier*") OR | TI("acidifier*") OR AB("acidifier*") OR |
|  | “Opiate Substitution Treatment”[MeSH] OR ((“opiate substitution” OR “opiate agonist” OR “opioid substitution” OR “opioid agonist”) AND (treatment or therapy)) OR | 'opiate substitution treatment'/exp OR “opioid agonist”:ab,ti OR “opiate substitution”:ab,ti OR | TITLE-ABS("opiate substitution") OR TITLE-ABS("opiate agonist") OR TITLE-ABS("opioid substitution") OR TITLE-ABS("opioid agonist") OR | TI("opiate substitution") OR AB(“opiate substitution”) OR | TI("opiate substitution") OR AB(“opiate substitution”) OR |
|  | “Medications for opioid use disorder”[tiab] OR MOUD[tiab] OR | “medications for opioid use disorder”:ab,ti OR MOUD:ab,ti OR | TITLE-ABS("Medications for opioid use disorder") OR TITLE-ABS("MOUD") OR | TI("Medications for opioid use disorder" OR “MOUD) OR AB("Medications for opioid use disorder" OR "MOUD") OR | TI("Medications for opioid use disorder" OR “MOUD) OR AB("Medications for opioid use disorder" OR "MOUD") OR |
|  | Methadone[tiab] OR | 'methadone treatment'/exp OR methadone:ab,ti OR | TITLE-ABS("Methadone") OR | TI("Methadone") OR AB("Methadone") OR | TI("Methadone") OR AB("Methadone") OR |
|  | Buprenorphine[tiab]) | Buprenorphine:ab,ti) | TITLE-ABS("Buprenorphine")) | TI("Buprenorphine") OR AB("Buprenorphine")) | TI("Buprenorphine") OR AB("Buprenorphine")) |
| Injecting-related infections | AND | AND | AND | AND | AND |
|  | (“injection-related infections”[tiab] OR “injection-related infection”[tiab] OR | ('injection site abscess'/exp OR “injection-related infections”:ab,ti OR “injection-related infection”:ab,ti OR | ((TITLE-ABS("injection-related infections") OR TITLE-ABS("injection-related infection") OR | (TI("injection-related infections" OR "injection-related infection") OR AB("injection-related infections" OR "injection-related infection") OR | (TI("injection-related infections" OR "injection-related infection") OR AB("injection-related infections" OR "injection-related infection") OR |
|  | “bacterial infection”[tiab] OR “bacterial infections”[tiab] OR | “bacterial infection”:ab,ti OR | TITLE-ABS("bacterial infection") OR TITLE-ABS("bacterial infections") OR | TI("bacterial infection" OR "bacterial infections") OR AB("bacterial infection" OR "bacterial infections") OR | TI("bacterial infection" OR "bacterial infections") OR AB("bacterial infection" OR "bacterial infections") OR |
|  | Bacteremia[MeSH] OR bacteremia[tiab] OR | 'bacteremia'/exp OR bacteremia:ab,ti OR | TITLE-ABS("bacteremia") OR | TI("bacteremia") OR AB("bacteremia") OR | TI("bacteremia") OR AB("bacteremia") OR |
|  | Fungemia[MeSH] OR | 'fungemia'/exp OR |  |  |  |
|  | Cellulitis[MeSH] OR cellulitis[tiab] OR | 'cellulitis'/exp OR cellulitis:ab,ti OR | TITLE-ABS("cellulitis") OR | TI("cellulitis") OR AB("cellulitis") OR | TI("cellulitis") OR AB("cellulitis") OR |
|  | Abscess[MeSH] OR abscess*[tiab] OR | 'abscess'/exp OR abscess*:ab,ti OR | TITLE-ABS("abscess*") OR | TI("abscess*") OR AB("abscess*") OR | TI("abscess*") OR AB("abscess*") OR |
|  | “skin infection”[tiab] OR “skin infections”[tiab] OR | “skin infection”:ab,ti OR “skin infections”:ab,ti OR | TITLE-ABS("skin infection") OR TITLE-ABS("skin infections") OR | TI("skin infection" OR "skin infections") OR AB("skin infection" OR "skin infections") OR | TI("skin infection" OR "skin infections") OR AB("skin infection" OR "skin infections") OR |
|  | “skin and soft tissue”[tiab] OR SSTI*[tiab] OR | “skin and soft tisuuse”:ab,ti OR SSTI*:ab,ti OR | TITLE-ABS("skin and soft tissue") OR TITLE-ABS("SSTI*") OR | TI("skin and soft tissue" OR "SSTI*") OR AB("skin and soft tissue" OR "SSTI*") OR | TI("skin and soft tissue" OR "SSTI*") OR AB("skin and soft tissue" OR "SSTI*") OR |
|  | Endocarditis[MeSH] OR endocarditis[tiab] OR | 'endocarditis'/exp OR endocarditis:ab,ti OR | TITLE-ABS("endocarditis") OR | TI(endocarditis) OR AB(endocarditis) OR | TI(endocarditis) OR AB(endocarditis) OR |
|  | Bone Diseases, Infectious[MeSH] OR |  |  |  |  |
|  | Osteomyelitis[MeSH] OR Osteomyelitis[tiab] OR | 'osteomyelitis'/exp OR osteomyelitis:ab,ti OR | TITLE-ABS("osteomyelitis") OR | TI(“osteomyelitis") OR AB(“osteomyelitis") OR | TI(“osteomyelitis") OR AB(“osteomyelitis") OR |
|  | “septic arthritis”[tiab] OR | “septic arthritis”:ab,ti OR | TITLE-ABS("septic arthritis")) | TI("septic arthritis") OR AB("septic arthritis")) | TI("septic arthritis") OR AB("septic arthritis")) |
|  | Central Nervous System Infections[MeSH] OR |  |  |  |  |
|  | Gram-Positive Bacterial Infections[MeSH] OR |  |  |  |  |
|  | Candidiasis[MeSH]) | 'candidiasis'/exp) |  |  |  |
| Social and structural determinants, or risk environment | AND | AND | AND | AND | AND |
|  | ("risk factor"[tiab] OR “risk factors”[tiab] OR | ('risk factor'/exp OR “risk factor”:ab,ti OR | (TITLE-ABS("risk factor") OR TITLE-ABS("risk factors") OR | (TI("risk factor” OR "risk factors") OR AB("risk factor” OR "risk factors") OR | (TI("risk factor” OR "risk factors") OR AB("risk factor” OR "risk factors") OR |
|  | correlate*[tiab] OR | correlate*:ab,ti OR | TITLE-ABS(correlate*) OR | TI("correlate*") OR AB("correlate*") OR | TI("correlate*") OR AB("correlate*") OR |
|  | determinant*[tiab] OR | determinant*:ab,ti OR | TITLE-ABS("determinant*") OR | TI("determinant*") OR AB("determinant*") OR | TI("determinant*") OR AB("determinant*") OR |
|  | environment*[tiab] OR | environment*:ab,ti OR | TITLE-ABS("environment*") OR | TI("environment*") OR AB("environment*") OR | TI("environment*") OR AB("environment*") OR |
|  | “social factors”[tiab] or “structural factors”[tiab] OR | 'social determinants of health'/exp OR “social factors”:ab,ti OR “structural factors”:ab,ti OR | TITLE-ABS("social factors") OR TITLE-ABS("structural factors") OR | TI("social factors" OR "structural factors") OR AB("social factors" OR "structural factors") OR | TI("social factors" OR "structural factors") OR AB("social factors" OR "structural factors") OR |
|  | Cohort*[tiab] OR | 'cohort analysis'/exp OR cohort*:ab,ti OR | TITLE-ABS(cohort*) OR | TI("Cohort*") OR AB("Cohort*") OR | TI("Cohort*") OR AB("Cohort*") OR |
|  | Longitudinal[tiab] OR | Longitudinal:ab,ti OR | TITLE-ABS("Longitudinal") OR | TI("Longitudinal") OR AB("Longitudinal") OR | TI("Longitudinal") OR AB("Longitudinal") OR |
|  | Prospective[tiab] OR retrospective[tiab] OR | Prospective:ab,ti OR retrospective:ab,ti OR | TITLE-ABS("Prospective") OR TITLE-ABS("retrospective") OR | TI("Prospective" OR "retrospective") OR AB("Prospective" OR "retrospective") OR | TI("Prospective" OR "retrospective") OR AB("Prospective" OR "retrospective") OR |
|  | Randomized[tiab] OR randomised[tiab] OR | Randomized:ab,ti OR randomised:ab,ti OR | TITLE-ABS("Randomized") OR TITLE-ABS("randomised") OR | TI("Randomized" OR "randomised") OR AB("Randomized" OR "randomised") OR | TI("Randomized" OR "randomised") OR AB("Randomized" OR "randomised") OR |
|  | Comparative[tiab] OR | Comparative:ab,ti OR | TITLE-ABS("Comparative") OR | TI("Comparative") OR AB("Comparative") OR | TI("Comparative") OR AB("Comparative") OR |
|  | Case-control[tiab] OR | Case-control:ab,ti OR | TITLE-ABS("Case-control") OR | TI("Case-control") OR AB("Case-control") OR | TI("Case-control") OR AB("Case-control") OR |
|  | Time-series[tiab] OR | 'time series analysis'/exp OR “time-series”:ab,ti OR | TITLE-ABS("Time-series") OR | TI("Time-series") OR AB("Time-series") OR | TI("Time-series") OR AB("Time-series") OR |
|  | Survey*[tiab] OR | Survey*:ab,ti OR | TITLE-ABS("Survey*") OR | TI("Survey*") OR AB("Survey*") OR | TI("Survey*") OR AB("Survey*") OR |
|  | Epidemiolog*[tiab] OR | Epidemiolog*:ab,ti OR | TITLE-ABS("Epidemiolog*") OR | TI("Epidemiolog*") OR AB("Epidemiolog*") OR | TI("Epidemiolog*") OR AB("Epidemiolog*") OR |
|  | Qualitative[tiab] OR | Qualitative:ab,ti OR | TITLE-ABS("Qualitative") OR | TI("Qualitative") OR AB("Qualitative") OR | TI("Qualitative") OR AB("Qualitative") OR |
|  | Interview[tiab] OR | 'interview'/exp OR interview:ab,ti OR | TITLE-ABS("Interview") OR | TI("Interview") OR AB("Interview") OR | TI("Interview") OR AB("Interview") OR |
|  | Ethnograph*[tiab] OR | 'ethnography'/exp OR ethnograph*:ab,ti OR | TITLE-ABS("Ethnograph*") OR | TI("Ethnograph*") OR AB("Ethnograph*") OR | TI("Ethnograph*") OR AB("Ethnograph*") OR |
|  | Mixed-methods[tiab] OR “mixed methods”[tiab] | Mixed-methods:ab,ti OR “mixed methods”:ab,ti OR | TITLE-ABS("Mixed-methods") OR TITLE-ABS("mixed methods") OR | TI("Mixed-methods" OR "mixed methods") OR AB("Mixed-methods" OR "mixed methods") OR | TI("Mixed-methods" OR "mixed methods") OR AB("Mixed-methods" OR "mixed methods") OR |
|  | gender[tiab] OR | Gender:ab,ti OR | TITLE-ABS(gender) OR | TI(gender) OR AB(gender) OR | TI(gender) OR AB(gender) OR |
|  | homeless*[tiab] OR | homeless*:ab,ti OR | TITLE-ABS(homeless*) OR | TI(homeless*) OR AB(homeless*) OR | TI(homeless*) OR AB(homeless*) OR |
|  | race[tiab] OR racism[tiab] OR | race:ab,ti OR racism;ab,ti OR | TITLE-ABS(race OR racism) OR | TI(race or racism) OR AB(race or racism) OR | TI(race or racism) OR AB(race or racism) OR |
|  | incarcerat*[tiab] OR prison*[tiab] OR criminal*[tiab] OR | incarcerat*:ab,ti OR prison*:ab,ti OR criminal*ab,ti OR | TITLE-ABS(incarcerat* OR prison* OR criminal*) OR | TI(incarcerat* OR prison* OR criminal*) OR AB(incarcerat* OR prison* OR criminal*) OR | TI(incarcerat* OR prison* OR criminal*) OR AB(incarcerat* OR prison* OR criminal*) OR |
|  | stigma*[tiab] OR discrimination[tiab] OR exclusion[tiab]) | stigma*ab,ti OR discrimination:ab,ti OR exclusion:ab,ti) | TITLE-ABS(stigma* OR discrimination OR exclusion*)) | TI(stigma* OR discrimination OR exclusion) OR AB(stigma* OR discrimination OR exclusion)) | TI(stigma* OR discrimination OR exclusion) OR AB(stigma* OR discrimination OR exclusion)) |
|  | *NOT (“case report”[Title]) NOT (“case series”[Title])* | NOT “case report”:ti NOT “case series”:ti NOT “rare case”:ti |  |  |  |
|  | Filter: 2000-Present | Filter:2000-2021 | Filter:2000-2021 | Limit to 2000-2021 | Limit to 2000-2021 |
|  |  | Filter: AND (**'article'**/it OR **'article in press'**/it OR **'conference abstract'**/it OR **'conference paper'**/it OR **'letter'**/it OR **'short survey'**/it)  [**To remove review articles] |  |  |  |
|  | February 18, 2021: 1,756 results  - Downloaded search results from PubMed as nbib file  - Imported into own folder in Zotero, confirmed 1756 items imported | February 18, 2021: 4,250 results  - Downloaded search results from Dalhousie EMBASE as RIS file | February 18, 2021: 835 results   - SCOPUS automatically exported to Zotero when I clicked Export as RIS | **February 18, 2021:** 300 results   - Persistent link to repeat search - Asked to send email with link to export results in RIS format | February 18, 2021: 92 results   - Asked to send email with link to export results in RIS format |
|  |  | Tried search again removing “preventive health services” and adding filters for “rare case” and filters to remove review articles & editorials:  3,506 results |  |  |  |

### Supplementary Table S2a. Critical appraisal of qualitative studies using the Mixed Methods Appraisal Tool (MMAT) for mixed studies systematic reviews.

|  |  | **Screening questions** | | **Qualitative studies** | | | | |
| --- | --- | --- | --- | --- | --- | --- | --- | --- |
|  | **Study** | **S1. Are there clear research questions?** | **S2. Do the collected data allow to address the research questions?** | **1. Is the qualitative approach appropriate to answer the research question?** | **2. Are the qualitative data collection methods adequate to address the research question?** | **3. Are the findings adequately derived from the data?** | **4. Is the interpretation of results sufficiently substantiated by data?** | **5. Is there coherence between qualitative data sources, collection, analysis and interpretation?** |
| 1 | Bearnot 2019 | Yes | Yes | Yes | Yes | Yes | Yes | Yes |
| 2 | Bearnot 2020 | Yes | Yes | Yes | Yes | Yes | Yes | Yes |
| 3 | Bodkin 2015 | Yes | Yes | Yes | Yes | Yes | Yes | Yes |
| 4 | Case 2008 | Yes | Yes | Yes | Yes | Yes | Yes | Yes |
| 5 | Colwill 2021 | Yes | Yes | Yes | Yes | Yes | Yes | Yes |
| 6 | Dunleavy 2019 | Yes | Yes | Yes | Yes | Yes | Yes | Yes |
| 7 | Epele 2002 | Yes | Yes | Yes | Yes | Yes | Yes | Yes |
| 8 | Gilbert 2019 | Yes | Yes | Yes | Yes | Yes | Yes | Yes |
| 9 | Harris RE 2018a | Yes | Yes | Yes | Yes | Yes | Yes | Yes |
| 10 | Harris RE 2018b | Yes | Yes | Yes | Yes | Yes | Yes | Yes |
| 11 | Harris M 2020a | Yes | Yes | Yes | Yes | Yes | Yes | Yes |
| 12 | Krüsi 2009 | Yes | Yes | Yes | Yes | Yes | Yes | Yes |
| 13 | Mars 2016 | Yes | Yes | Yes | Yes | Yes | Yes | Yes |
| 14 | McNeil 2014 | Yes | Yes | Yes | Yes | Yes | Yes | Yes |
| 15 | Meyer 2020 | Yes | Yes | Yes | Yes | Yes | Yes | Yes |
| 16 | Paquette 2018 | Yes | Yes | Yes | Yes | Yes | Yes | Yes |
| 17 | Pollini 2021 | Yes | Yes | Yes | Yes | Yes | Yes | Yes |
| 18 | Sheard 2008 | Yes | Yes | Yes | Yes | Yes | Yes | Yes |
| 19 | Small 2008 | Yes | Yes | Yes | Yes | Yes | Yes | Yes |

### Supplementary Table S2b. Critical appraisal of mixed methods studies using the Mixed Methods Appraisal Tool (MMAT) for mixed studies systematic reviews.

|  |  | **Screening questions** | | **Mixed methods studies** | | | | |
| --- | --- | --- | --- | --- | --- | --- | --- | --- |
|  | **Study** | **S1. Are there clear research questions?** | **S2. Do the collected data allow to address the research questions?** | **1. Is there an adequate rationale for using a mixed methods design to address the research question?** | **2. Are the different components of the study effectively integrated to answer the research question?** | **3. Are the outputs of the integration of qualitative and quantitative components adequately interpreted?** | **4. Are divergences and inconsistencies between quantitative and qualitative results adequately addressed?** | **5. Do the different components of the study adhere to the quality criteria of each tradition of the methods involved?** |
| 1 | Bourgois 2006 | Yes | Yes | Yes | Yes | Yes | Yes | Yes |
| 2 | Harris M 2019 | Yes | Yes | Yes | Yes | Yes | Yes | Yes |
| 3 | Harris M 2020b | Yes | Yes | Yes | Yes | Yes | Yes | Yes |
| 4 | Jafari 2015 | Yes | Yes | Yes | Yes | No | No | No |
| 5 | Phillips 2013 | Yes | Yes | Yes | Yes | Yes | Yes | Yes |
| 6 | Pollini 2010 | Yes | Yes | Yes | Yes | Yes | Yes | Yes |
| 7 | Summers 2018 | Yes | Yes | Yes | Yes | Yes | Yes | Yes |
